# Incorporating family disease history and controlling case-control imbalance for population based genetic association studies

**DOI:** 10.1101/2021.07.04.21259997

**Authors:** Yongwen Zhuang, Brooke N Wolford, Kisung Nam, Wenjian Bi, Wei Zhou, Cristen J Willer, Bhramar Mukherjee, Seunggeun Lee

## Abstract

In the genome-wide association analysis of population-based biobanks, most diseases have low prevalence, which results in low detection power. One approach to tackle the problem is using family disease history, yet existing methods are unable to address type I error inflation induced by increased correlation of phenotypes among closely related samples, as well as unbalanced phenotypic distribution. We propose a new method for genetic association test with family disease history, TAPE (mixed-model-based Test with Adjusted Phenotype and Empirical saddlepoint approximation), which controls for increased phenotype correlation by adopting a two-variance-component mixed model, accounts for case-control imbalance by using empirical saddlepoint approximation, and is flexible to incorporate any existing adjusted phenotypes such as phenotypes from the LT-FH method. We show through simulation studies and analysis of UK-Biobank data of white British samples and the Korean Genome and Epidemiology Study (KoGES) of Korean samples that the proposed method is robust and yields better calibration compared to existing methods while gaining power for detection of variant-phenotype associations.

## Introduction

Genome-wide and phenome-wide studies are facilitated by the recent development of large-scale biobanks such as the UK Biobank (UKB)^1^, BioBank Japan (BBJ)^2^ and the Korean Genome and Epidemiology Study (KoGES)^3^. Individuals in the biobanks are samples from a target population and large numbers of phenotypes are collected for each individual, which allows phenome-wide scans. However, challenges remain to gain enough power to identify associated variants, especially for binary traits with a low prevalence.

One promising approach to improve detection power is using family disease history to infer risk of diseases of unaffected individuals. For family-based cohorts with partially-missing genotypes, association test power can be improved by using pedigree information^4-7^. The GWAX method first demonstrated that with completely-missing family genotypes, unaffected individuals with family disease history can be used as proxy-cases to find genetic associations^8^. The LT-FH method^9^ further increases association power by estimating a liability of disease conditional on the observed phenotypes and family disease history, which differentiate the disease risks among the proxy cases.

Despite the progress, several important limitations remain. First, when samples are related, the increased correlation among the inferred risks (**Figure S1**) can lead to type I error inflation. Hujoel et. al showed that since samples with close relatedness such as sibling pairs tend to have highly correlated GWAX or LT-FH phenotypes due to nearly identical family disease history, GWAX and LT-FH suffered poor calibration compared to GWAS ^9^. Thus, the usage of the existing methods should be restricted to testing unrelated individuals only, which can reduce power. Second, with unbalanced case-control ratios, the distributions of inferred risks are still unbalanced, hence testing for association using linear mixed model (LMM) can yield inflated type I error rates. For example, diseases such as Parkinson’s disease have low prevalence in UK-Biobank, which leads to a small number of cases and proxy cases (i.e. controls with non-zero inferred disease risk) in GWAX and a relatively low posterior liability conditioning on family history in LT-FH (**Figures S2 and S3**). Since the gaussian approximation does not perform well in this setting, LMMs can yield inflated type I error rates. Currently no method exists to handle situations of this kind.

We propose a new method for genetic association test with family disease history, TAPE (mixed-model-based Test with Adjusted Phenotype and Empirical saddlepoint approximation), which controls for increased phenotype correlation and case-control imbalance. In standard mixed model methods, only a dense genetic relatedness matrix is used as the variance component. TAPE uses a sparse kinship matrix as an additional variance component to further account for the increased correlation among phenotypes in closely related individuals. In addition, to adjust for case-control imbalance, TAPE uses empirical saddlepoint approximation under a linear mixed model^10-12^.

The TAPE method takes a three-step framework: (0) infer the disease risk for all individuals in the analysis based on the original case-control status and family disease history to be used as phenotype; (1) fit two variance components null linear mixed model to obtain parameter estimates; (2) test for genetic association using score test with empirical saddlepoint approximation. The analytical framework of TAPE also has the flexibility to use adjusted phenotypes generated from other approaches as the outcome variable. In addition to deriving adjusted phenotype using the weighted proportion of kinship coefficients of affected relatives (denoted as TAPE-WP), we also used the inferred disease risk in LT-FH as an adjusted phenotype and run Steps 1 and 2 mentioned above (denoted as TAPE-LTFH).

We show through simulation studies and analysis of UK Biobank that the proposed method is robust and yields better calibration compared to existing methods while gaining power for detection of variant-phenotype associations. We applied TAPE-WP and TAPE-LTFH to 10 binary traits in UK Biobank among 408,898 white British samples with imputed genotypes and parental disease information. TAPE-WP identified 663 genome-wide significant clumped variants, among which 33 were with MAF<1%, and TAPE-LTFH identified a total of 726 genome-wide significant clumped, including 63 clumped variants with MAF < 1%. We also analyzed two binary phenotypes in the KoGES data with 72,298 samples and identified 29 genome-wide significant clumped variants in total using TAPE.

## MATERIAL AND METHODS

### Overview of methods

The TAPE method takes a three-step framework (**Figure 1**): (0) infer the disease risk for all individuals in the analysis based on the original case-control status and family disease history to be used as phenotype; (1) fit two variance components null linear mixed model to obtain parameter estimates; (2) test for genetic association using score test with empirical saddlepoint approximation.

**Figure 1.**
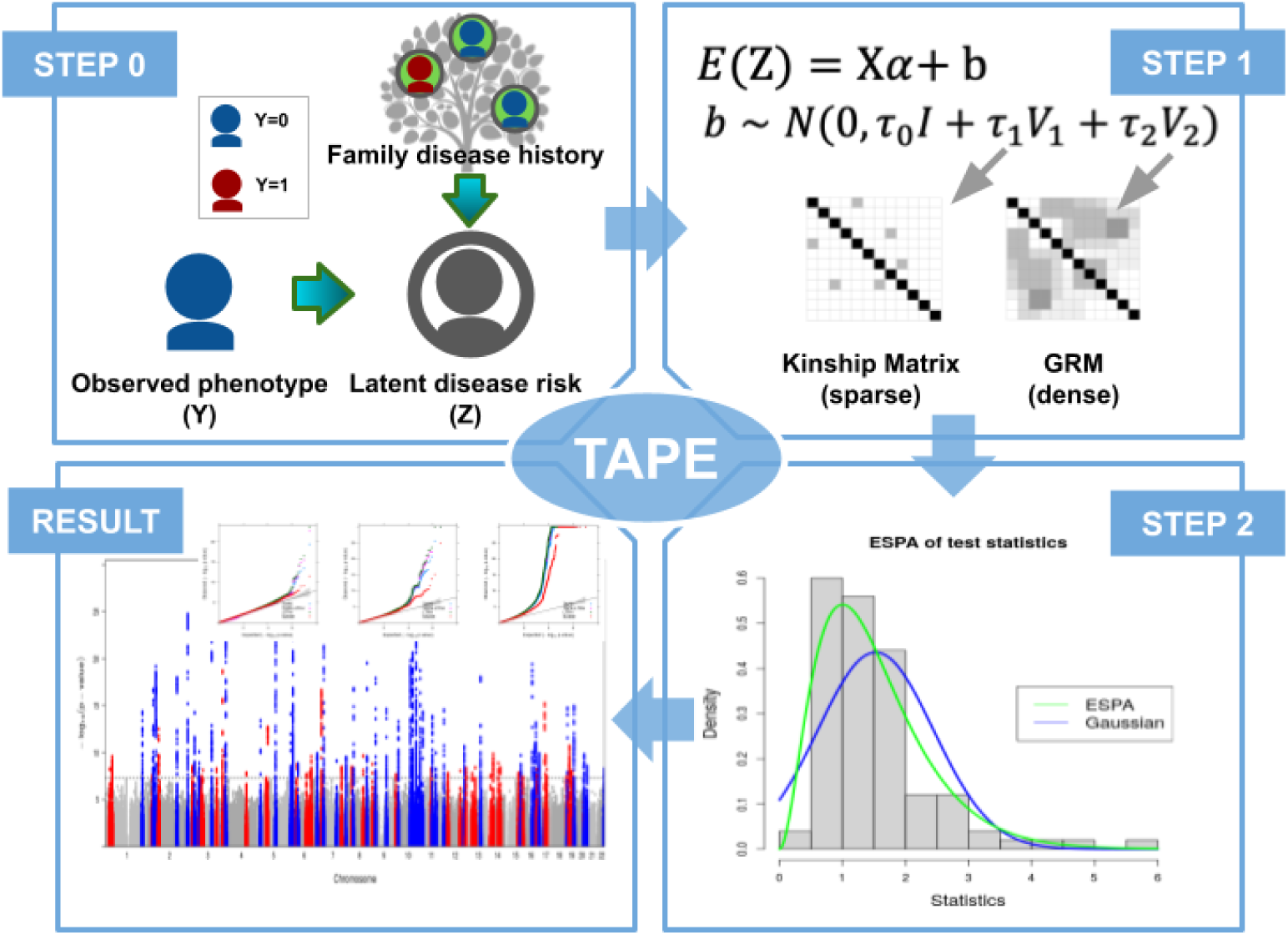
Analytical framework of TAPE. In Step 0, latent disease risk of individuals is estimated from observed phenotypes and family disease history using a weighted proportion of the affected close relatives to the individual. In Step 1, a null linear mixed model is fit with covariates and two random effects with the sparse kinship matrix and the dense genetic relatedness matrix (GRM) as covariance structures. In Step 2, p-values score test is performed for each genetic variant using empirical saddlepoint approximation.

In Step 0, the phenotypes are adjusted using inferred risk of individuals. TAPE-WP uses a weighted proportion of the affected close relatives to the control, which can be viewed as an extension of the GWAX method^8^ to further differentiate disease risk of controls based on family disease history configurations. TAPE-LTFH uses the liability of diseases generated from the existing LT-FH method as the adjusted phenotypes.

In Step 1, we fit the null linear mixed model to estimate model parameters. Fixed effects of the null model include covariates such as demographic information and principal components. Sample relatedness is accounted for by random effects in the model. We include two random effects, the first uses the sparse kinship matrix as covariance structure similar to that in fastGWA^26^, and the second uses the dense genetic relatedness matrix (GRM). These two variance components can capture both increased correlation in phenotypes due to phenotype adjustment procedure and distance genetic relatedness among individuals. To make the method scalable, average information restricted maximum likelihood (AI-REML)^15^, with preconditioned conjugate gradient (PCG) method^16^ similar to that used in BOLT-LMM^17^ and SAIGE, is used. The computation complexity is *𝒪*(*B*(*M*_*GRM*+_*C*_*sparse*_)*N*^1.5^) where *B* is the number of iterations until convergence, *C*_*sparse*_ is the number of non-zero elements in the sparse kinship matrix, *M*_*GRM*_ is the number of variants included in the GRM construction, *N* is the sample size, and the number of iterations for PCG method is assumed to be *𝒪*(*N*^0.5^). We use raw genotypes as input and calculate GRM in runtime, yielding a reduced memory usage of *M*_*GRM*_*N*/4 bytes compared to methods that facilitate a precomputed GRM, which has memory usage of *fN*^2^ bytes where *f* denotes memory size for a floating number.

In Step 2, score test statistic is calculated for each genetic variant against the adjusted phenotype. Since the Gaussian approximation does not perform well at the tails of the test statistic distribution especially when the case-control ratio is unbalanced and MAF of the variant is low, we approximate the distribution by empirical saddlepoint approximation^11^, which uses empirically estimated cumulant generating function (CGF) to calculate p-value. The empirical saddlepoint approximation is utilized when the test statistic exceeds two standard deviations of the mean. Time complexity for this step is *𝒪*(*MN*).

### Phenotype adjustment

We first introduce the proposed phenotype adjustment procedure (Step 0) in TAPE. For TAPE-WP, we assume a sample of *N* individuals where each individual has 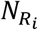 relatives with phenotypic information (*i* ∈ {1, …, *N*}), *F*_*ij*_ denotes the kinship coefficient between individual *i* and relative 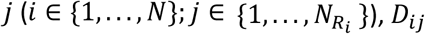 denotes the phenotype of relative *j* of individual *i, Y* is an *N*-vector of observed binary phenotypes. The adjusted quantitative phenotype for individual *i, Z*_*i*_, is expressed as:

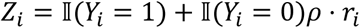

where 𝕀(·) denotes indicator function, *ρ* is a pre-specified constant indicating the increase in latent disease risk, and 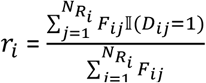. If *Y_i_* = 0 and all 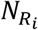 relatives of the *i*th individual are cases, the latent disease risk is *Z*_*i*_ = *ρ*. For the analysis in this paper, we assume that latent risk of such individual is 0.5 (i.e., *ρ* = 0.5). In addition, the phenotype adjustment procedure can be adapted to include information other than family disease status that is potentially indicative of latent disease risk. See **Supplementary Note 1 and 2** for details.

For TAPE-LTFH, we use the posterior mean genetic liability from the LT-FH method proposed by Hujoel et al. ^9^ as the outcome variable in the analysis, which is computed conditioning on test samples’ binary phenotypes and available disease status of parents and siblings through Monte Carlo integration.

### Linear mixed model (LMM) for adjusted phenotype

We denote *X*_*i*_ as a (p+1)-vector of covariates with the intercept, and *G*_*i*_ as the allele counts for the variant to be tested. We consider the following linear model:

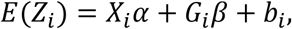

where *α* is a (*p*+1)-vector of fixed effect coefficients, *β* is a genetic effect coefficient, and *b*_*i*_ the random effect term for the *i*th individual with *b* = (*b*_1_, …, *b*_*N*_)^*T*^. We as sum e the r ando m effec t to follow a multivariate Gaussian distribution 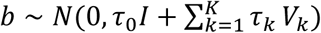, where *τ*_0_ is the variance component parameter for a noise term. Parameters for other variance components are denoted as *τ*_*k*_, and *V*_*k*_ are pre-specified *N* × *N* correlation matrices.

To better capture phenotype correlation, we use a variance component of sparse kinship in addition to the commonly used genetic relationship matrix (GRM), i.e., *K* = 2 and *Σ* = *τ*_0_*I*+*τ*_1_ *V*_1+_*τ*_2_*V*_2_, where *V*_1_ is a sparse matrix of the estimated kinship coefficients after thresholding, and *V*_2_ is GRM computed from genetic variants. The inclusion of the sparse kinship matrix as an additional variance component can be justified by the observation that the phenotype adjustment using family disease information increases the concordance among related individuals. For example, the adjusted phenotype for a control sibling pair would be identical as they share the same parental disease status (**Figure S1**). Such phenotypic concordance is not sufficiently captured by GRM alone and can lead to mis-calibration as pointed out by Hujoel et al.^9^. It is also shown that incorporating pedigree structure as a variance component in linear mixed models improves association outcomes^13, 14^.

### Parameter estimation for the null model

In Step 1, we fit a linear mixed model under the null hypothesis of no genetic effects and estimate its parameters. The null model can be represented as

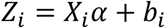

Treating the adjusted phenotype *Z* as a quantitative trait, the log likelihood of (*α, τ*) with random effect *b* integrated out in REML is

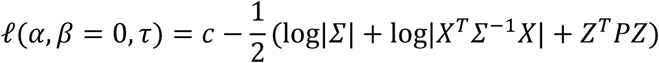

where *c* is a constant,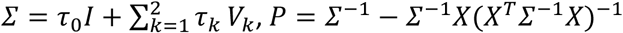. Model parameters (*α, β, τ*) are estimated iteratively with a working model 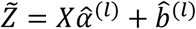 for iteration *l*. Let 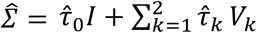 be the working variance matrix. The first derivatives of *l*(*α, β* = 0, *τ*) with respect to *τ* are:

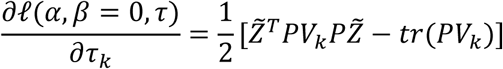

For each iteration, variance components 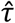 are updated using AI-REML algorithm^15^, in which the Hessian is approximated by an average information matrix *AI* with its entries expressed as:

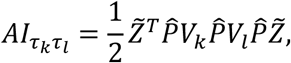

where 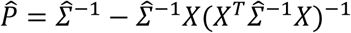. Then the variance component parameters are updated by 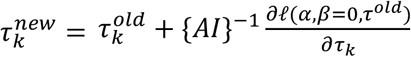.

Both the first derivative and the approximated second derivative involves matrix inverse of 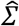, which can be computationally heavy when *N* is large. To reduce the computational burden, the PCG method^16^ with Jacobi preconditioner is adopted, which avoids directly calculating matrix inverse by finding solutions of linear systems and involves only matrix multiplication. Since 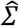 is a linear combination involving two components *V*_1_ and *V*_2_ in our setting, matrix multiplication with regard to the two parts can be calculated separately. For *V*_1_ which is a sparse matrix representing close relatedness up to the third degree, the computation cost is further lowered by scanning through the non-zero elements of *V*_1_ only. For *V*_2_ which represents genetic relatedness, we improve the memory usage by calculating its elements in runtime instead of using a pre-computed *N* × *N* GRM matrix. Thus, the overall time complexity for null model estimation is reduced from *𝒪*(*N*^3^) to *𝒪*(*B*(*M*_*GRM*+_*C*_*sparse*_)*N*^1.5^), where *B* is the number of iterations until the algorithm reaches convergence, *C*_*sparse*_ is the number of non-zero elements of sparse kinship matrix, *M*_*GRM*_ is the number of variants included in the GRM construction. Here we assume that PCG algorithm has complexity *𝒪*(*N*^0.5^).^17^

To avoid double-fitting the candidate variant in the model and GRM, leave-one-chromosome-out (LOCO) scheme was implemented in both the null model parameter estimation step and the score statistic calculation step of the proposed method.

### Single variant association test with empirical SPA

In Step 2, we use the score test to calculate the p-value for association of each genetic variant against the adjusted phenotype. The score statistic for testing the null hypothesis *H*_0_: *β*_*j*_ = 0 for variant *j* is 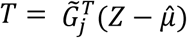, where 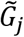 is an N-vector of covariate-adjusted genotypes. Under the null hypothesis, the variance of the statistic is 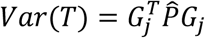. For computational efficiency, *Var*(*T*) can be approximated using 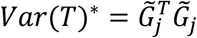 combined with a calibration factor 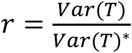 estimated using a subset of SNP data. The variance-adjusted statistic after calibration is 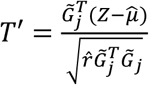. For the proposed method, 30 SNPs were used to obtain the estimated calibration factor 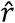.

When Z is unbalanced and a variant has low minor allele count, the distribution of *T′* deviates from the Gaussian distribution especially at the tails, thus the usual test of the score statistic against a Gaussian distribution can result in type I error inflation. Saddlepoint approximation is shown to improve over normal approximation in such conditions by utilizing the entire cumulant-generating function (CGF)^20, 21^. Fixing 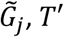 can be viewed as a weighted sum of residuals 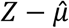, yet the adjusted phenotype *Z* has an intractable distribution which makes it impossible to derive the explicit cumulant-generating function (CGF).

Alternatively, we use the empirical version of saddlepoint approximation^10, 11^ as a nonparametric estimator for the distribution of the test statistic. The empirical CGF approach has been utilized in methods such as SPACox^12^ for an altered version of the test statistic and was shown to provide calibrated p-values. The empirical estimator for CGF of *T′* is 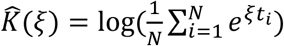 where *t*_*i*_ is the residual of the *i*th individual from Step1. The empirical approximation of the first and second derivative are 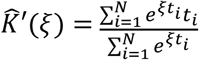 and 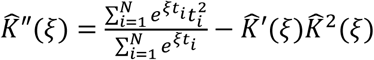 respectively. Suppose 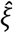 is a value satisfying the equation 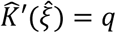, the p-value can be calculated by the following formula^22^

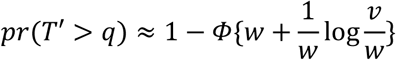

where 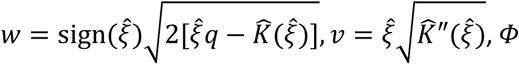, is the cumulative distribution function of the standard normal distribution.

### Simulation studies

We performed simulation studies to evaluate type I error rate and power of the proposed method. To simulate a population of size *N* with *p*_*r*_% relatedness, we set *p*_*r*_*N*/100 to be related individuals with a specified relatedness structure, and the rest 1 − *p*_*r*_*N*/100 to be independent individuals. We considered two types of relatedness structure for the simulated population. The first one consists of 5,000 independent individuals and 2,500 sibling pairs (*p*_*r*_ = 50%). The second one is a mixture of independent individuals and families with 8 members in each family. The pedigree for the 8-member family was shown in **Figure S5**. To obtain a sample size of 10,000 with 50% related individuals, 625 families were simulated with the 8-member pedigree, while the rest 5,000 are independent individuals. In the sibling-pair scenario, the simulation process proceeds as follows: First, sequences of *M* variants for both parents of each sample individual were simulated independently with pre-specified MAFs. Genotypes for *N* sample individuals (offsprings) were then generated using *N*_*G*_ = 2*N* par ental gen otypes. Bin ary phe notypes for sample individuals and parents were simulated from *Bernoulli*(*μ*_*i*_) with *μ*_*i*_ from a logistic mixed model

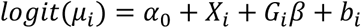

where for individual *i* (*i* = 1, …, 3*N*), *X*_*i*_ is a covariate randomly sampled from *Normal*(0,1), *G*_*i*_ is the genotypes of the M variants, *α*_0_ is the intercept determined by prevalence *k, β* is a vector of log odds ratio of genetic effects, and *b*_*i*_ is a random effect with underlying distribution *Normal* (0, *τK*) depending on the true underlying kinship coefficient matrix *K*. Given the kinship coefficient *φ*_*ij*_ between individual *i* and individual *j*, the value for an element in *K* is *K*_*ij*_ = 2*φ*_*ij*_.

Simulation results for TAPE-WP and TAPE-LTFH were compared with two other methods: (1) GWAS with original binary phenotypes by SAIGE^19^; (2) Original LT-FH method that uses BOLT-LMM with LT-FH phenotypes^9^ (hereafter denoted as LT-FH), which is shown to increase association power over GWAX ^8^.

Type I error rates were evaluated at significant level *α* = 5 × 10^−8^ for simulated data sets with 10^9^ independent null SNPs, and sample size 10,000 at case-control ratio of 1:99, 5:95 and 10:90. Phenotypes were generated given *τ* = 1, corresponding to liability-scale heritability 0.23^19^. To investigate type I error rates by MAF, SNPs were generated with MAF 0.001, 0.01 and 0.1 respectively for each simulated data set.

Power of the tests was assessed using simulated data sets with 10,000 individuals and 100,000 variants with MAF 0.1 for each setting with 1% variants selected as causal variants. We calculated both the average *χ*^2^ statistics for causal SNPs and the empirical power at empirical *α* level from SAIGE, LT-FH, TAPE-LTFH and TAPE-WP. Genetic effect sizes ranged from 0.4 to 2.3 and three case-control ratio settings were considered, i.e., 1:99, 5:95, and 10:90. We generated 100 replications for each simulation scenario.

### Computation time

Computation time was evaluated with *M* = 100,000 variants and sample size *N* ranging from 10,000 to 408,898 sampled from white British individuals in UK Biobank data for Type II diabetes (case:control=1:20). Projected time for the analysis of 21 million variants with MAF ≥ 0.01 % was calculated based on the evaluation results. Two other methods were evaluated in addition to TAPE-WP: analysis of binary phenotypes by SAIGE and analysis of LT-FH phenotypes by BOLT-LMM. All evaluations were computed on an Intel(R) Xeon(R) Gold 6152 CPU.

### UK Biobank data

Over 21 million genetic variants imputed from the Haplotype Reference Consortium (HRC)^23^ and with minor allele frequency (MAF) ≥ 0.01% were used for the association analysis among a sample population of 408,898 white British individuals. NCBI Build 37/UCSC hg19 was adopted for genomic coordinates. A total of 10 binary traits with available parental disease status were analyzed, where the binary traits for genotyped individuals were defined by the PheWAS codes^19^ aggregated from ICD9 and ICD10 codes in the UK Biobank. Parental phenotypes were extracted from data fields for self-reported paternal and maternal illness. We included sex, age and first 10 principal components as covariates to adjust for. GRM was constructed using 93,511 genotyped variants suggested by UK Biobank^19, 24^. Kinship coefficients were estimated using the KING software^25^, and the sparse kinship matrix was constructed using those with estimated kinship no larger than third-degree relatedness. Calibration of the testing method was evaluated by the attenuation ratio obtained from stratified LD score regression (LDSC). The attenuation ratio is defined as (LDSC intercept -1)/(average *χ*^2^-1), with smaller values indicating better control of false positives.

### The KoGES data

For the association analysis among a sample population of 72,298 Korean individuals, over 8 million genetic variants were imputed from 1,000 Genome project phase 3 + Korean reference genome (397 samples) and with minor allele frequency (MAF) > 1%^3^. Two binary traits (diabetes and gastric cancer) with different case-control ratios were analyzed. Phenotypes for both genotyped individuals and their relatives are self-reported survey data. We adjusted for sex, age, first 10 principal components, and 34 indicator variables of batch information (cohort × collection year). GRM was constructed using 327,540 genotyped variants. The sparse GRM was constructed using SAIGE with pairwise relatedness coefficients larger than 0.1.

## RESULTS

### Simulation study results

#### Type I error and Power

Type I error rates were evaluated at genome-wide *α* = 5 × 10^−8^ with sample size of 10,000 and case-control ratio ranging from 1:99 to 10:90. For each case-control ratio setting, two sets of genotype data with 10^9^ independent variants were generated with MAF of 0.1, 0.01 and 0.001 respectively. We first simulated a population consisting of 2,500 pairs of siblings and 5,000 independent individuals (**Table 1)**. The empirical type I error rates of LT-FH were significantly inflated under more unbalanced case-control ratio and lower MAF, while results from TAPE-WP and SAIGE were well calibrated. TAPE-LTFH also yielded better controlled type I error rates than that of LT-FH, especially when the case-control ratio is more unbalanced (1:99), since the additional variance component in the mixed model further accounts for the phenotypic concordance among sibling pairs with same family disease history, and the empirical saddlepoint approximation better approximates the distribution of test statistics. Further, we evaluated type I error rates with a more complex relatedness structure, i.e., a population consisting of 625 8-member families and 5,000 independent individuals (**Table 1)**. Inflated type I error rates were observed in results from LT-FH but with lower magnitude compared to the previous setting. TAPE-LTFH had slightly inflated type I error rates. One explanation is that LT-FH phenotypes are less concordant in the latter setting since there is a smaller number of individuals sharing identical family history under a more complicated pedigree. On the other hand, type I error rates from TAPE and SAIGE were relatively well controlled with a slight deflation.

**Table 1.** Empirical type I error rates for TAPE-WP, TAPE-LTFH, LT-FH and SAIGE, estimated using 10^9^ independent SNPs and a sample size of 10,000 (*α* = 5 × 10^−8^). Two types of population structure were considered: 1) Sample consists of 2,500 pairs of siblings and 5,000 independent individuals; 2) Sample consists of 625 8-member families and 5,000 independent individuals.

| Case:Control | MAF | TAPE-WP | TAPE-LTFH | LTFH | SAIGE |
| --- | --- | --- | --- | --- | --- |
| *2,500 pairs of siblings and 5,000 independent individuals* |  |  |  |  |  |
| 1:99 | 0.001 | 4.977e-08 | 1.019e-07 | 5.928e-06 | 4.418e-08 |
| 5:95 | 0.001 | 5.115e-08 | 8.275e-08 | 1.252e-06 | 4.368e-08 |
| 10:90 | 0.001 | 5.476e-08 | 7.452e-08 | 5.489e-07 | 4.641e-08 |
| 1:99 | 0.01 | 5.455e-08 | 1.069e-07 | 1.409e-07 | 3.963e-08 |
| 5:95 | 0.01 | 5.143e-08 | 1.158e-07 | 1.940e-07 | 4.341e-08 |
| 10:90 | 0.01 | 5.459e-08 | 9.086e-08 | 1.141e-07 | 4.980e-08 |
| 1:99 | 0.10 | 5.007e-08 | 1.275e-07 | 1.500e-07 | 3.964e-08 |
| 5:95 | 0.10 | 5.213e-08 | 1.639e-07 | 1.238e-07 | 4.355e-08 |
| 10:90 | 0.10 | 6.416e-08 | 7.782e-08 | 7.232e-08 | 4.650e-08 |
| *625 8-member families and 5,000 independent individuals* |  |  |  |  |  |
| 1:99 | 0.001 | 3.329e-08 | 9.028e-08 | 4.446e-06 | 3.832e-08 |
| 5:95 | 0.001 | 3.051e-08 | 6.563e-08 | 8.171e-07 | 4.245e-08 |
| 10:90 | 0.001 | 2.967e-08 | 5.145e-08 | 3.751e-07 | 4.721e-08 |
| 1:99 | 0.01 | 3.742e-08 | 9.792e-08 | 4.818e-07 | 4.547e-08 |
| 5:95 | 0.01 | 3.156e-08 | 7.906e-08 | 1.463e-07 | 4.311e-08 |
| 10:90 | 0.01 | 2.978e-08 | 6.215e-08 | 8.811e-08 | 4.324e-08 |
| 1:99 | 0.10 | 3.113e-08 | 7.730e-08 | 1.000e-07 | 3.895e-08 |
| 5:95 | 0.10 | 3.050e-08 | 7.983e-08 | 6.025e-08 | 4.232e-08 |
| 10:90 | 0.10 | 3.163e-08 | 6.372e-08 | 5.857e-08 | 4.546e-08 |

One of the important features of TAPE is the use of a kinship matrix in addition to (dense) GRM to account for increased correlation among phenotypes. Two additional analyses were performed to investigate the influence of no kinship variance component (TAPE-nok) and mis-specified kinship matrix (TAPE-misk) on calibration of TAPE under the 8-member family pedigree scenario. For TAPE-nok, the sparse kinship matrix was not included as an LMM variance component and inflated empirical type I error was observed (**Figure S4)**. For TAPE-misk, the true kinship matrix of an 8-member family pedigree was replaced with a slightly mis-specified one (**Figure S5)** in step 0 and step 1. The empirical type I error of TAPE-misk was similar to that of TAPE. The results indicated that the impact of a slightly mis-specified kinship matrix was negligible, while the inclusion of the kinship matrix as a variance component is crucial in controlling type I error rate when family information is incorporated into the analysis.

To assess empirical power, we compared the average *χ*^2^ statistics of causal SNPs (**Figure 2**) and the proportion of causal SNPs significant at empirical *α* level (**Figure S6**) for simulated data sets with sample size 10,000 under different genetic effects and case-control ratio. For each data set, 100,000 independent variants with MAF 0.1 were simulated in which 1% were causal, and we generated 100 data sets for each setting. TAPE-WP and TAPE-LTFH achieves greater detection power over SAIGE, with a 21.0% and 26.5% average increase in average *χ*^2^ statistics, and a 18.3% and 22.4% average increase in proportion of causal SNPs detected, respectively. LT-FH also had increased *χ*^2^ over SAIGE by 27.5% and had a 22.1% average increase in detection rate, but it suffered from type I error inflation especially when analyzing related samples.

**Figure 2.**
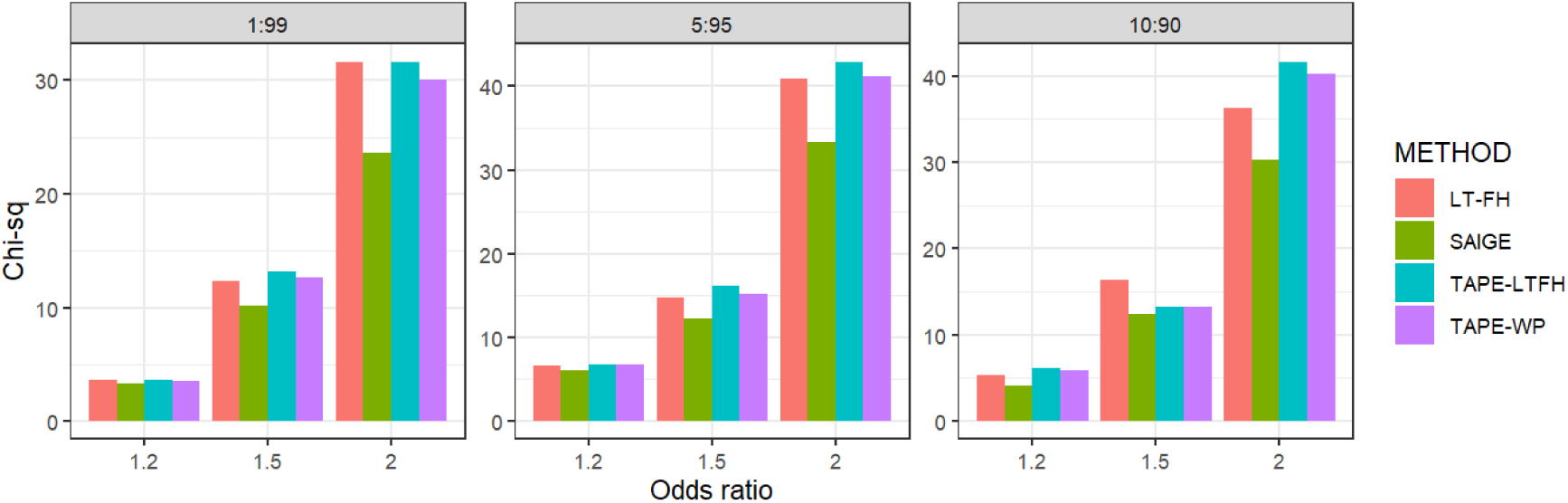
Average *χ*^2^ values of causal variants with sample size N=10,000 (5,000 independent individuals and 2,500 pairs of siblings), comparing TAPE-WP, TAPE-LTFH, LT-FH and SAIGE. For each dataset, 100,000 independent variants were simulated and 1% variants were selected as causal variants with 4 different effect sizes. A total of 100 datasets were generated to calculate average *χ*^2^ values. MAFs of variants were 0.1.

To investigate how more complex relatedness structures will influence simulation results, we further simulated a population in which related individuals form families with 8 members (**Figure S5)**. TAPE-WP achieves greater detection power over SAIGE GWAS results, with a 24.1% average increase in average *χ*^2^ statistics and a 30.6% average increase in proportion of causal SNPs detected. TAPE-LTFH has higher overall power than both LT-FH and SAIGE under such relatedness structure (**Figure S6**), with a 27.5% average increase in average *χ*^2^ statistics and a 40.8% average increase in proportion of significant SNPs detected over SAIGE, whereas for LT-FH the increase is 25.6% for average *χ*^2^ statistics and 36.6% for proportion of significant SNPs detected.

In general, TAPE-WP and TAPE-LTFH yielded well-controlled type I error rate even when case-control ratio is unbalanced, which makes the incorporation of family disease information in genetic association test feasible in the presence of sample relatedness and gains detection power. In addition, TAPE-LTFH achieved higher detection power than LT-FH, especially under the simulation scenario with more complex relatedness structure.

#### Computation Time

Computation time was evaluated using randomly selected samples from 408,898 white British individuals in UK Biobank data for Type II diabetes (case:control=1:20) with *M* = 100,000 variants. Projected computation time for 21 million variants with MAF ≥ 0.01% was estimated and plotted on log10 scale against sample size varying from 10,000 to 408,898 (**Figure S7**). Computation time for TAPE-LTFH is similar to that for TAPE-WP and is therefore omitted in the plot. A break-down of run time for null model estimation and p-value calculation is presented in **Table S3**. Since TAPE fits the model with two variance components and uses ESPA in p-value calculation, which requires additional computation, TAPE was slower than SAIGE and LT-FH. Overall, TAPE is scalable to analyze biobank size data. For genome-wide analysis of testing 21 million variants, TAPE required 16 CPU hours with 40,000 samples and 284 CPU hours with 408,898 samples.

### Analysis of binary traits in biobank data

We analyzed 10 binary disease outcomes with available parental disease status in the UK Biobank^1^. The binary traits were defined by the PheWAS codes^19^ aggregated from ICD codes in the UK Biobank dataset, with case-control ratio ranging from 1:4 to 1:406 among 408,898 white British individuals (**Table 2**). We tested over 21 million variants imputed from the Haplotype Reference Consortium (HRC)^23^ with minor allele frequency (MAF) ≥ 0.01%. Sex, age and first 10 principal components were included in the analysis model. GRM was constructed using 93,511 genotyped variants with high quality. Kinship coefficients were estimated using the KING software^25^, and the sparse kinship matrix was constructed using those with estimated kinship no larger than third-degree relatedness.

**Table 2.**
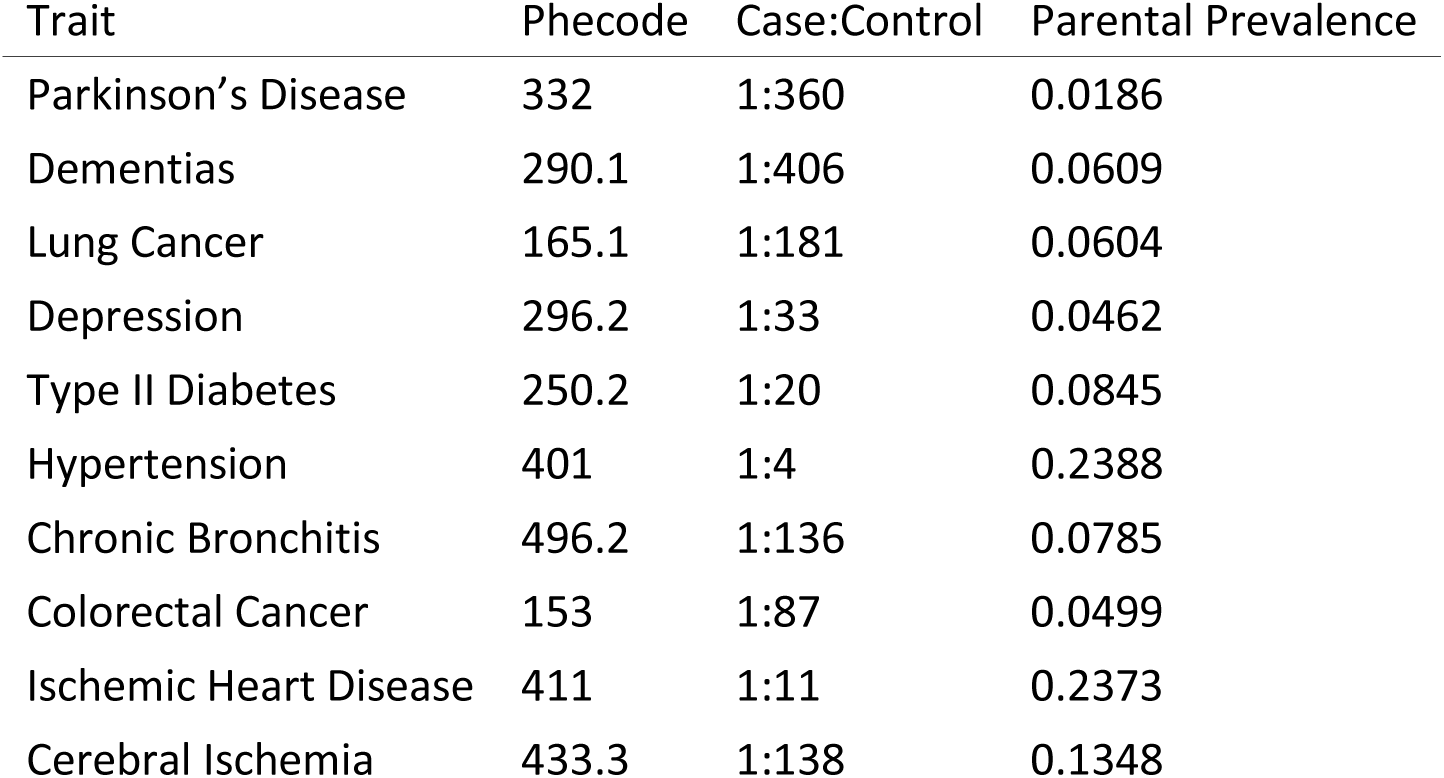
Summary of 10 traits in UK Biobank

| Trait | Phecode | Case:Control | Parental Prevalence |
| --- | --- | --- | --- |
| Parkinson's Disease | 332 | 1:360 | 0.0186 |
| Dementias | 290.1 | 1:406 | 0.0609 |
| Lung Cancer | 165.1 | 1:181 | 0.0604 |
| Depression | 296.2 | 1:33 | 0.0462 |
| Type II Diabetes | 250.2 | 1:20 | 0.0845 |
| Hypertension | 401 | 1:4 | 0.2388 |
| Chronic Bronchitis | 496.2 | 1:136 | 0.0785 |
| Colorectal Cancer | 153 | 1:87 | 0.0499 |
| Ischemic Heart Disease | 411 | 1:11 | 0.2373 |
| Cerebral Ischemia | 433.3 | 1:138 | 0.1348 |

**Figures 3 and 4** present Manhattan plots and Q-Q plots stratified by MAF categories for two phenotypes with different case-control ratio: Type II diabetes (case-control ratio 1:20), and Parkinson’s disease (case-control ratio 1:350). Plots for all 10 diseases in the analysis are shown in **Figures S8 and S9**. For diseases such as Parkinson’s Disease (**Figure 4**), the observed quantile distribution of -log10(p) corresponding to SNPs with MAF < 0.01 for LT-FH method in the QQ plot curve off in the middle of the graph, indicating potential type I error inflation due to unaccounted-for relatedness structure. Similar problematic patterns can also be found in LT-FH QQ plots for lung cancer, depression, chronic bronchitis, colorectal cancer and cerebral ischemia (**Supplementary Figure 8**), but not in plots for TAPE-WP and TAPE-LTFH.

**Figure 3.**
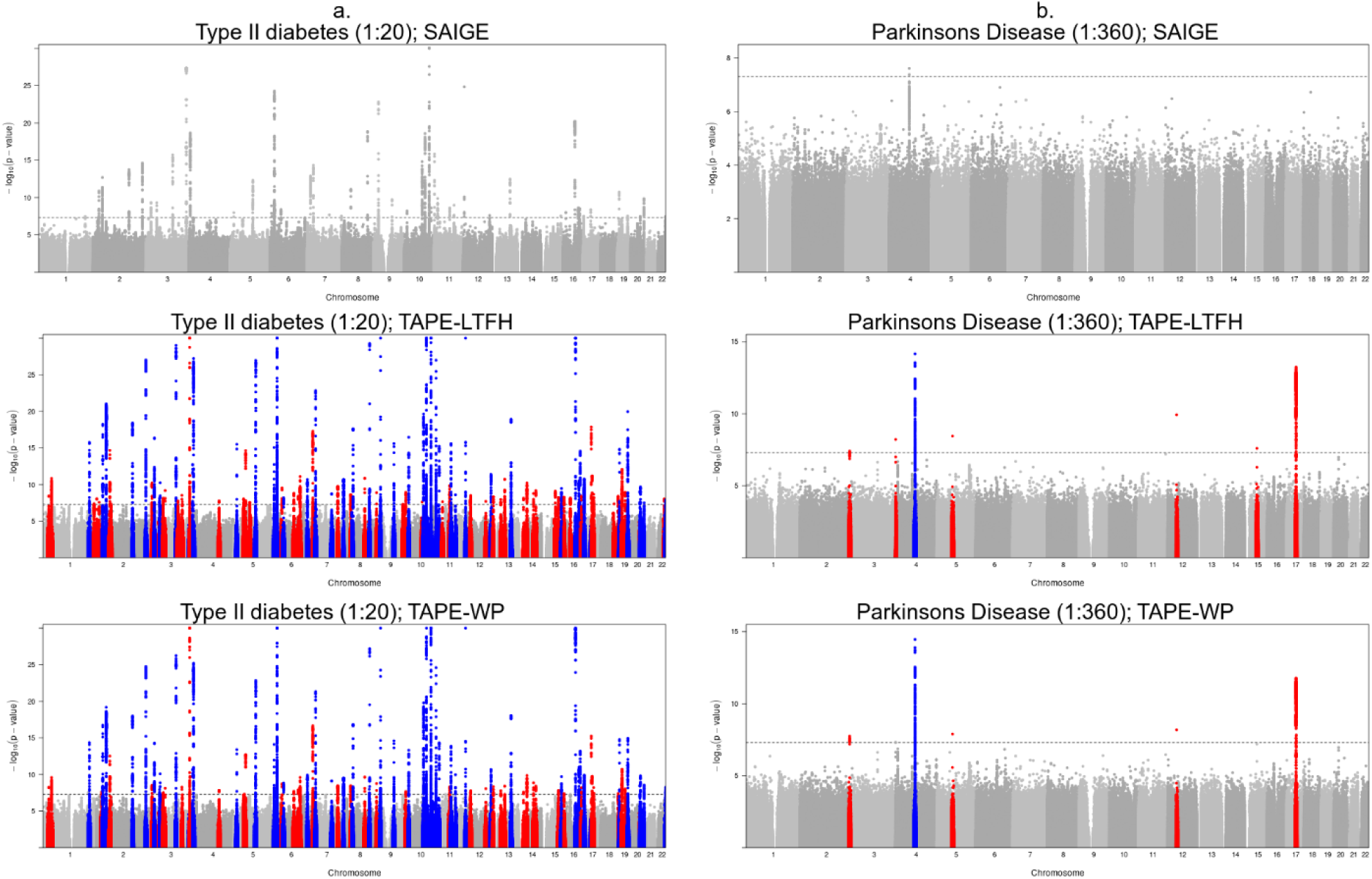
Manhattan plot for the UK Biobank association test results from SAIGE (first row), TAPE-LTFH (second row) and TAPE-WP (third row) among white British (N=408,898). Left: Type II diabetes (Phecode 250.2); Right: Parkinson’s disease (Phecode 332). For plots from TAPE-WP and TAPE-LTFH, red marks clumped significant variants that were not detected by SAIGE; blue marks clumped significant variants detected by both TAPE-WP and SAIGE. Significant clumped variants are identified using a window width of 5Mb and a linkage disequilibrium threshold of 0.1.

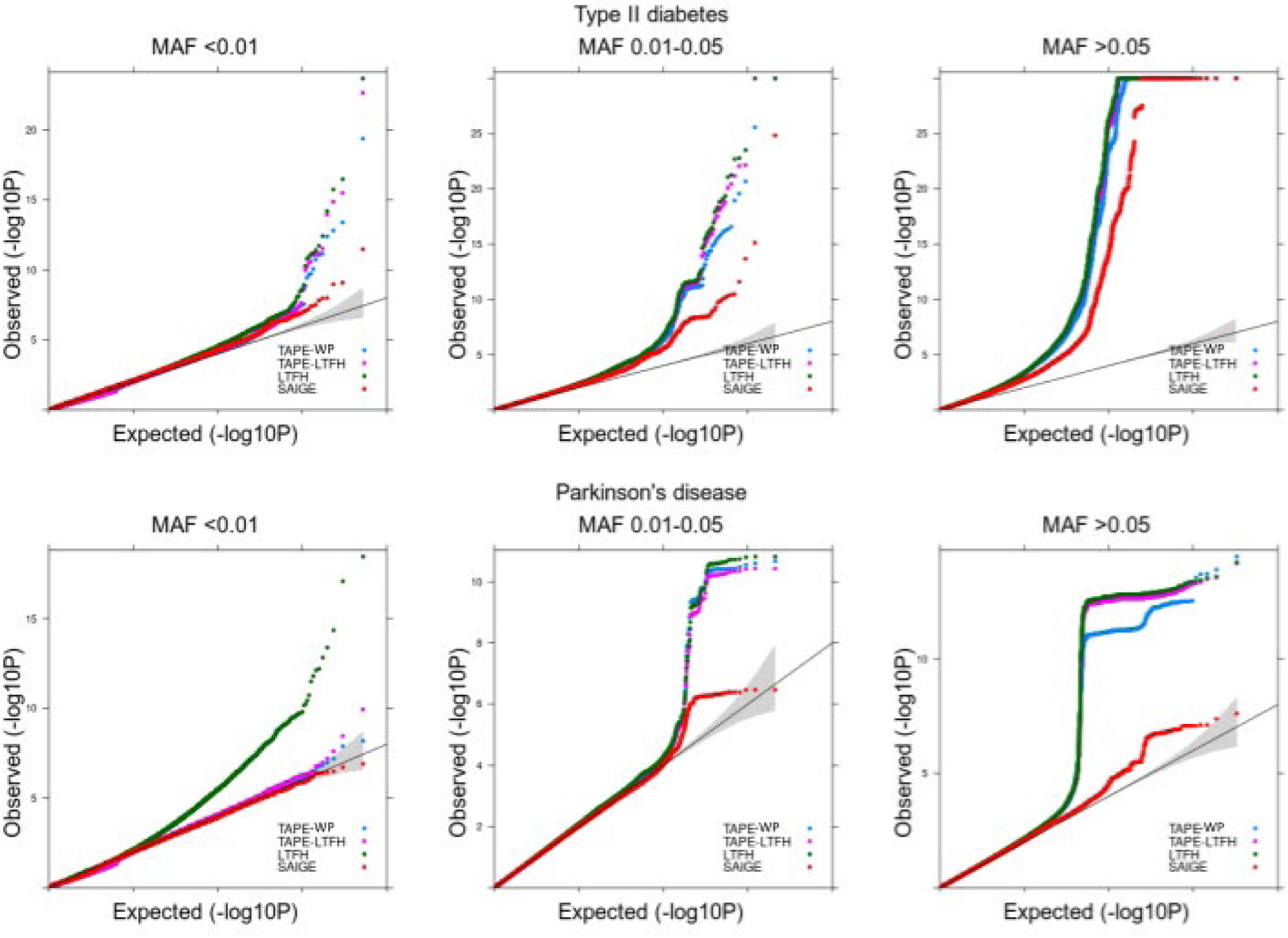

To assess the calibration of testing methods, we performed stratified LD score regression with the baselineLD model to obtain the attenuation ratios^28^ (**Table S2**). For traits with more unbalanced case-control, TAPE-WP consistently yields relatively lower attenuation ratios than TAPE-LTFH, while LT-FH generates the highest attenuation ratio, indicating poor calibration. For example, the average attenuation ratio for type II diabetes (case:control=1:20) is 0.110, 0.120 and 0.142 for TAPE-WP, TAPE-LTFH and LT-FH respectively; for Parkinson’s disease (case:control=1:360), the average attenuation ratio is 0.125, 0.222 and 0.462 for TAPE-WP, TAPE-LTFH and LT-FH respectively. Since we used all the individuals regardless of relatedness, the observation supports the previously reported result that LT-FH suffers poor calibration in related samples due to concordance between phenotypes from closely related samples such as sibling pairs^9^. On the contrary, TAPE-WP is able to generate better calibrated results under such situations, followed by TAPE-LTFH. Due to the above-mentioned potential type I error inflation of LT-FH method for samples with relatedness, we only compare the proposed method with SAIGE, which is also capable of handling related samples. **Table S1** lists the number of significant variants and significant clumped variants at *α* = 5 × 10^−8^ detected by TAPE-WP, TAPE-LTFH and SAIGE. Significant clumped variants were further identified by clumping genome-wide significant variants with 5Mb window size and linkage disequilibrium threshold *r*^2^ = 0.1 using PLINK software^27^. TAPE-WP identified 84 more genome-wide significant clumped variants than SAIGE for Type II diabetes, and 5 more for Parkinson’s disease. For TAPE-LTFH, a total of 111 more genome-wide significant clumped variants were identified for Type II diabetes, and 7 more for Parkinson’s disease as compared to SAIGE. For all 10 diseases analyzed, a total of 663 genome-wide significant clumped variants were identified by TAPE-WP, including 33 clumped variants with MAF < 1%; whilst a total of 344 clumped variants were identified by SAIGE, of which 25 were with MAF < 1%. For TAPE-LTFH, a total of 726 genome-wide significant clumped variants were identified, including 63 clumped variants with MAF < 1%.

For additional analysis, we applied TAPE-WP, TAPE-LTFH and SAIGE to two binary phenotypes for 72,298 individuals with family disease history in the KoGES data and analyzed 8 million variants. Disease prevalence among sample individuals and their relatives is shown in **Table S4**. For diabetes (case:control=1:12), TAPE-WP identified 14 more genome-wide significant clumped variants than SAIGE, while TAPE-LTFH identified 15 more than SAIGE. For gastric cancer (case:control=1:191), both TAPE-WP and TAPE-LTFH identified 3 genome-wide significant clumped variants (rs760077, rs35972942, rs2978977) while no variants were genome-wide significant by SAIGE. The three clumped variants have been previously reported to be associated with gastric cancer among Chinese or Japanese population ^29-31^, but not among Korean samples. Manhattan plots and Q-Q plots are presented in **Figures S10 and S11**.

## DISCUSSION

We propose a robust method that incorporates family disease information for genetic association test while accounting for case-control unbalance and close relatedness in the population. Samples in biobanks usually follow cohort study design, which can have small number of cases compared to traditional case-control study design, especially for rare phenotypes. Previous studies have shown that additional information from family disease history can help improve test power in such cases, yet challenges remain (1) to control for type I error inflation induced by increased correlation of phenotypes among related individuals after incorporating family disease history; and (2) to account for unbalanced distribution of phenotypes after being adjusted by family disease information. TAPE-WP and TAPE-LTFH (hereafter denoted as the TAPE framework) use both a dense genetic relatedness matrix and a sparse matrix for close relatedness as variance components in the linear mixed model framework to account for sample relatedness and family-history-induced correlations. Empirical saddlepoint approximation of the score test statistic distribution is adopted to control for type I error inflation under unbalanced phenotypic distribution. Optimization strategies such as PCG for computing components with matrix inversion, and runtime GRM calculation from raw genotypes were implemented to improve computation efficiency and reduce memory usage.

The TAPE framework incorporates family disease history by adjusting the phenotypes of control samples with a product of *ρ* and *r*, where *ρ* indicates the increase in latent disease risk among controls given all relatives of the individual are cases, and *r* represents a weighted proportion of diseased relatives of the individual. We assumed a constant *ρ* = 0.5 for the analysis in this paper, and we expect more accurate estimates of this value to yield better performance in capturing potential disease risk among controls with family disease history, which might be a direction for future exploration. For example, *ρ* can be estimated in a similar way as genetic nurturing effects for different phenotypes respectively under a family analysis framework where genotypes for relatives are avaiable^16^, thus enabling different level of contribution from relatives to the individual’s latent risk for different diseases.

For the null model, both sparse estimated kinship matrix and GRM are included in the TAPE framework as variance components to account for the potential phenotypic concordance. The use of two or more variance components in mixed model has been shown to better control for test statistics inflation and improves association power as well as prediction accuracy in standard GWAS and family studies ^32, 33^, yet we are not aware of existing methods that apply more than one variance components to mixed model while incorporating family disease history. From simulation studies we show that the absence of kinship matrix in variance components leads to inflated type I error rates of association test results. This result echoes previous findings from LT-FH that the inclusion of family disease history can lead to miscalibration under a single-variance-component mixed model due to similar family history for closely related individuals such as sibling pairs^9^, and indicates a possible solution to control for phenotypic correlation introduced by incorporating family disease information. When estimating variance parameters, the TAPE framework improves computation efficiency by applying PCG algorithm on top of the sparse estimated kinship matrix and the dense GRM, where sparsity of the estimated kinship matrix is ascertained by proper thresholding.

The analytical framework of TAPE allows for flexible choice of outcome variables. For example, TAPE-LTFH uses LT-FH phenotypes in the proposed two-variance-component mixed model. We show by simulation studies that TAPE-LTFH can better control for type I error inflation than LT-FH and achieves higher power. It remains a future work to better capture latent risk while accounting for phenotypic concordance to further improve association power using external information such as family disease history. The TAPE framework currently supports analysis of binary phenotypes, and has the potential to be extended to ordinal phenotypes, such as categorical diagnoses of mental disorders, for more powerful association test in a broader context.

We also note several limitations of our proposed method. First, the potential difference in the phenotype classification for genotyped individuals and their relatives is not accounted for in the TAPE framework. For example, phenotypes of genotyped individuals in UKB dataset were defined using the PheWAS codes aggregated from ICD9 and ICD10 codes, whereas parental phenotypes were extracted from self-reported surveys. The different phenotype classification standard may induce bias in the adjusted phenotype after incorporating family disease history. The second limitation lies in the modeling assumption of infinitesimal genetic effects, i.e., the effect size of each variant follows a standard Normal distribution, which may yield less detection power when the assumption does not match the true underlying genetic architecture.

Despite the above-mentioned limitations, the TAPE framework is the only existing approach that incorporates family disease history while handling related samples and phenotype unbalance. With the increasing accessibility to large-scale biobank data with population relatedness and family disease history information, our proposed method is expected to contribute to improving detection power for genetic association studies, especially for late-onset diseases that are underrepresented in the sample cohorts.

## Data Availability

The authors confirm that the data supporting the findings of this study are available within the article and its supplementary materials.

## SUPPLEMENTAL DATA

Supplemental data include two notes, twelve figures and three tables.

## DECLARATION OF INTERESTS

The authors declare no competing interests.

## ACKNOWLEDGMENTS

This research was supported by NIH grants R01-LM012535 and R01-HG008773 (Y.Z.), and Brain Pool Plus (BP+, Brain Pool+) Program through the National Research Foundation of Korea (NRF) funded by the Ministry of Science and ICT (2020H1D3A2A03100666, S.L). UK Biobank data were accessed under the accession number UKB: 45227.

## AUTHOR CONTRIBUTIONS

Y.Z., C.J.W and S.L. designed the experiments. Y.Z., B.W. and S.L. analyzed the UK Biobank data. K.N. and S.L. analyzed the KoGES data. Y.Z implemented the program with help from W.B and W.Z. Y.Z wrote the manuscript with input and critical feedback from all authors.

## WEB RESOURCES

TAPE (version 0.3.0): https://github.com/styvon/TAPE.

TAPE summary association statistics for 10 phenotypes in UKB data and 2 phenotypes in KoGES data: https://github.com/styvon/TAPE/blob/main/vignettes/biobank_results.md

BOLT-LMM (version 2.3.4): https://data.broadinstitute.org/alkesgroup/BOLT-LMM.

LT-FH (version 2): https://alkesgroup.broadinstitute.org/UKBB/LTFH.

SAIGE (version 0.44.3): https://github.com/weizhouUMICH/SAIGE.

KING (version 2.2.4): https://www.kingrelatedness.com/.

PLINK (version 2.00): https://www.cog-genomics.org/plink2.

## DATA AND CODE AVAILABILITY

The code generated during this study are available at https://github.com/styvon/TAPE

## Notes

### Competing Interest Statement

The authors have declared no competing interest.

### Funding Statement

This research was supported by NIH grants R01-HG008773 (Y.Z.), and Brain Pool Plus (BP+, Brain Pool+) Program through the National Research Foundation of Korea (NRF) funded by the Ministry of Science and ICT (2020H1D3A2A03100666, S.L). UK Biobank data were accessed under the accession number UKB: 45227.

### Author Declarations

This work does not need any approval of the IRB.

