## Supplementary Materials for "Incorporating family disease history and controlling case-control imbalance for population based genetic association studies"

**Supplementary Note**

**1. Derivation of adjusted phenotype for TAPE-WP**

Here we consider a sample of size $N$. Suppose each individual has $N_{R_i}$ relatives with known phenotype. Let $F_{ij}$ denote the kinship coefficient between individual $i$ and its $j$'th relative, and let $D_{ij}$ denote the phenotype for the $j$'th relative of individual $i$. Denote $Y_i$ as the observed phenotype of individual $i$, $Y'_i$ as a latent phenotype indicating whether the individual will have the disease in the future. $Z_i = Pr(Y'_i = 1|Y_i, F_{i\cdot}, D_{i\cdot})$ as the adjusted phenotype indicating latent disease risk given the observed phenotype $Y_i$ and relative information. With the assumption that $Pr(Y'_i = 1|Y_i = 1, F_{i\cdot}, D_{i\cdot}) = 1$, we get the following equation:

$$Z_i \quad = \mathbb{I}(Y_i = 1) + \mathbb{I}(Y_i = 0)Pr(Y'_i = 1|Y_i = 0, F_{i\cdot}, D_{i\cdot})$$

We form an estimator of $Pr(Y'_i = 1|Y_i = 0, F_{i\cdot}, D_{i\cdot})$ by

$$\widehat{Pr}(Y'_i = 1|Y_i = 0, F_{i\cdot}, D_{i\cdot}) = \rho \cdot r_i$$

where $\rho$ is a pre-specified constant indicating the increase in latent disease risk among controls given all $N_{R_i}$ relatives of an individual are cases. For the analysis in this paper $\rho = 0.5$ was used. We further define $r_i = \frac{\sum_{j=1}^{N_{R_i}} F_{ij}\mathbb{I}(D_{ij}=1)}{\sum_{j=1}^{N_{R_i}} F_{ij}}$, which is a measure for the proportion of affected relatives weighted by kinship coefficient.

As an illustrative example, consider a simple setting where each individual has phenotypic information of one parent (kinship coefficient is 0.25), i.e., $N_{R_i} = 2$ and $F_{i1} = 0.25$, $(i = 1, \ldots, N)$. If an unaffected individual $i$ has an affected parent, then $r_i = 1$, the adjusted phenotype $Z_i = 0.5$, which is equivalent to GWAX[1].

### 2. Extension of adjusted phenotype to include age information

The phenotype adjustment step of TAPE can be easily applied to account for information indicative of latent disease risk among controls other than family disease status. One potentially useful information is age. The aging process plays a crucial part in late-onset diseases (LODs), influencing both the missing heritability and case-control imbalance in observational studies, which adds to the difficulty in obtaining sufficient power in testing the association between disease and genetic variants. Several theories were established to examine the mechanism of LODs. The mutation accumulation model states that genetic products generated since birth gradually accumulate as one ages, which can eventually lead to the manifestation of disease at an older age[21]. The latent damage model of antagonistic pleiotropy explains the disease effect in elder age as a result of the late effect from a mutation occurred earlier in life[22]. Patterns of LOD development are further explored using computer simulations, which shows that for diseases such as Alzheimer's Disease and type II diabetes, there is a rapid change in the difference of effective variant minor allele frequency between cases and controls along with age. Study results also showed that the average polygenetic risk among unaffected individuals decreases with older age[23]. Recall from previous section the formula to estimate $Pr(Y'_i = 1|Y_i = 0, F_{i\cdot}, D_{i\cdot})$:

$$\widehat{Pr}(Y'_i = 1|Y_i = 0, F_{i\cdot}, D_{i\cdot}) = \rho \cdot r_i$$

where $\rho$ indicates the increase in latent disease risk among controls given all $N_R$ relatives of an individual are cases and was previously set to be constant across all controls. To capture the potential influence of age on a control's disease risk, we can use survival curve as a surrogate of polygenic risk at a certain age and construct $\rho$ as a function of $A_i$, age of individual $i$:

$$\rho_i = \rho_0 \hat{S}(A_i = a)$$

with $\hat{S}(\cdot)$ being an estimator for survival function $S(\cdot)$ and $\rho_0$ a pre-defined scaling parameter. Several different approaches exist to estimate the survival function, including Life Table and Kaplan-Meier approach, but is not the focus of this paper.

**3. Supplementary Figures**

**Supplementary Figure 1.** Illustration of phenotypic concordance between related individuals. Suppose both parents are affected with a disease with a relatively low prevalence in a family consisting of two parents (1,2) and two offsprings (3,4), then the estimated latent risks for 3 and 4 are both Z=0.5 considering only parental disease history. Since 3 and 4 share the same family disease status, they get identical value of estimated latent risk as adjusted phenotype in the TAPE framework, while for non-related individuals the probability of getting the same family disease status is lower and the gap enlarges with a lower disease prevalence. As a result, the concordance of adjusted phenotype is higher among related individuals compared to independent individuals.

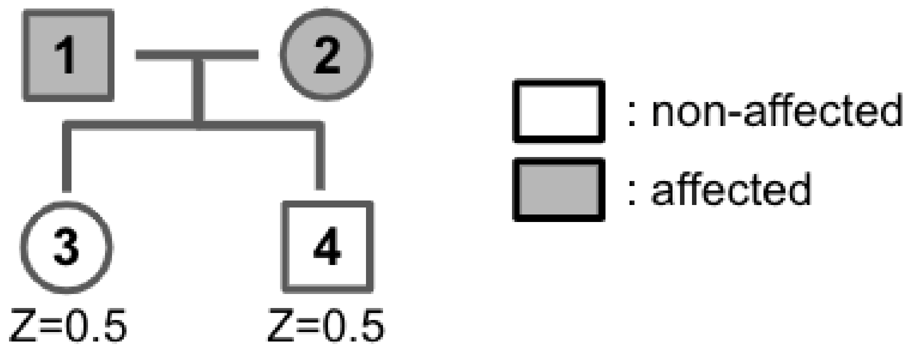

**Supplementary Figure 2.** Skewed distribution of inferred disease risk after incorporating family disease history for 10 UK Biobank binary traits with sample size 408,898, comparing TAPE-WP and LT-FH. For each method, the quantitative inferred disease risk is divided into 5 bins of equal size with width $(max - min/6)$.

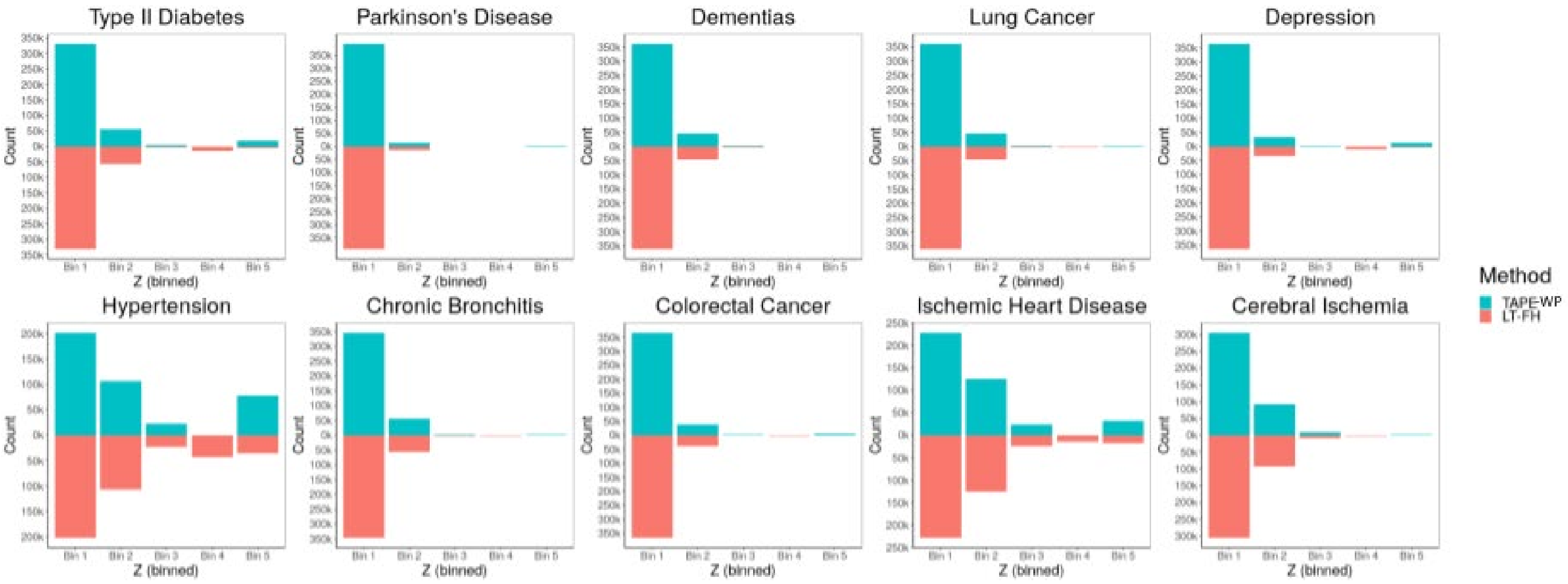

**Supplementary Figure 3.** Skewed distribution of inferred disease risk in simulation data with sample size 10,000 and case-control ratios ranging from 1:99 to 10:90, comparing GWAX and LT-FH. For each method, the quantitative inferred disease risk is divided into 5 bins of equal size with width $(max - min/6)$.

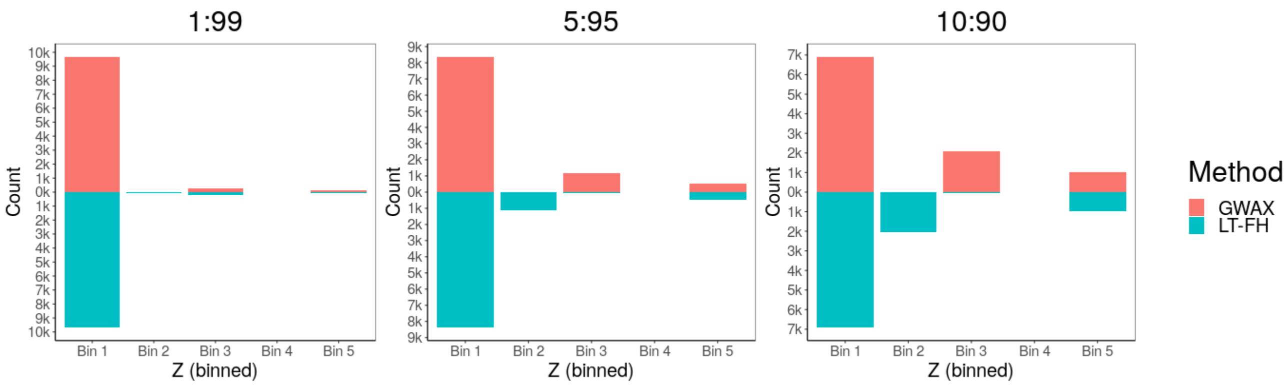


**Supplementary Figure 4.** Q-Q plot for additional simulation analyses, comparing TAPE-WP, TAPE-misk, TAPE-nok.

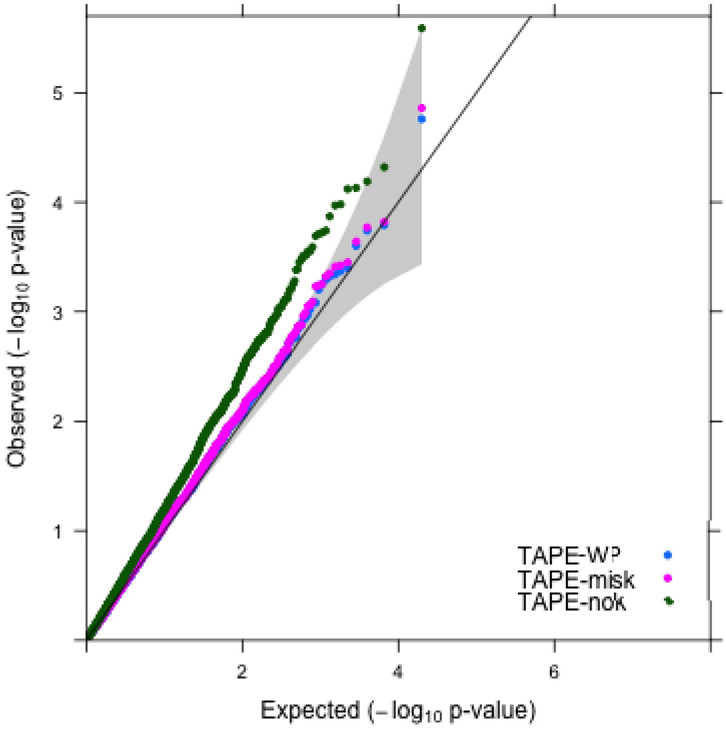

**Supplementary Figure 5.** Pedigree for the 8-member family. **a**: True relatedness structure. The family consists of individuals 3-10, but excludes founders 1 and 2 in the dotted box. **b**: Mis-specified relatedness structure. Siblings 3 and 4 are treated as independent.

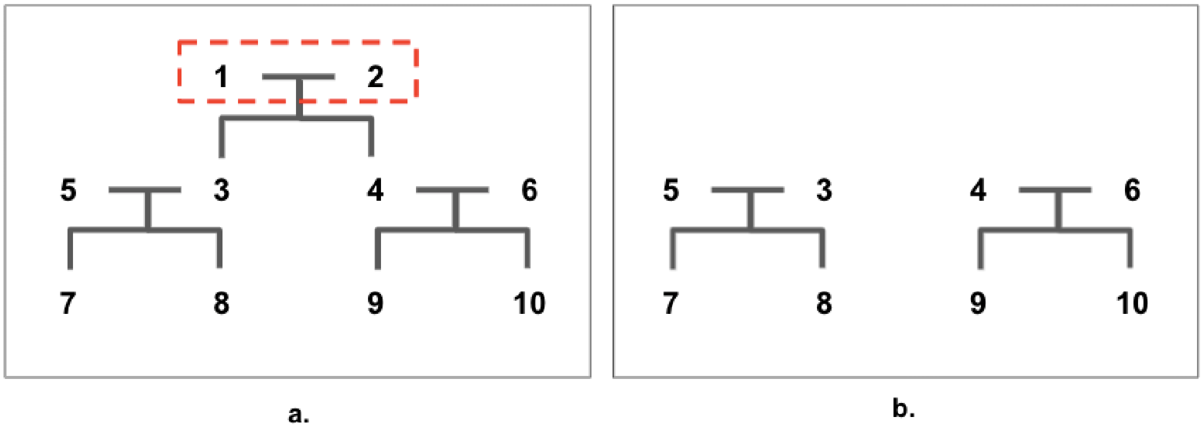

**Supplementary Figure 6.** Simulation results for empirical power for causal variants with sample size N=10000, comparing TAPE-WP, TAPE-LTFH, LT-FH and SAIGE using empirical $\alpha$. **a**: Mixture population containing 5000 independent individuals and 2500 pairs of siblings; **b**: Mixture population containing 5000 independent individuals and 625 families with 8 members each. For each set, a total of 100,000 independent variants were simulated and 1% variants were selected as causal variants with 4 different effect sizes. A total of 100 datasets were generated to calculate empirical power.

a.

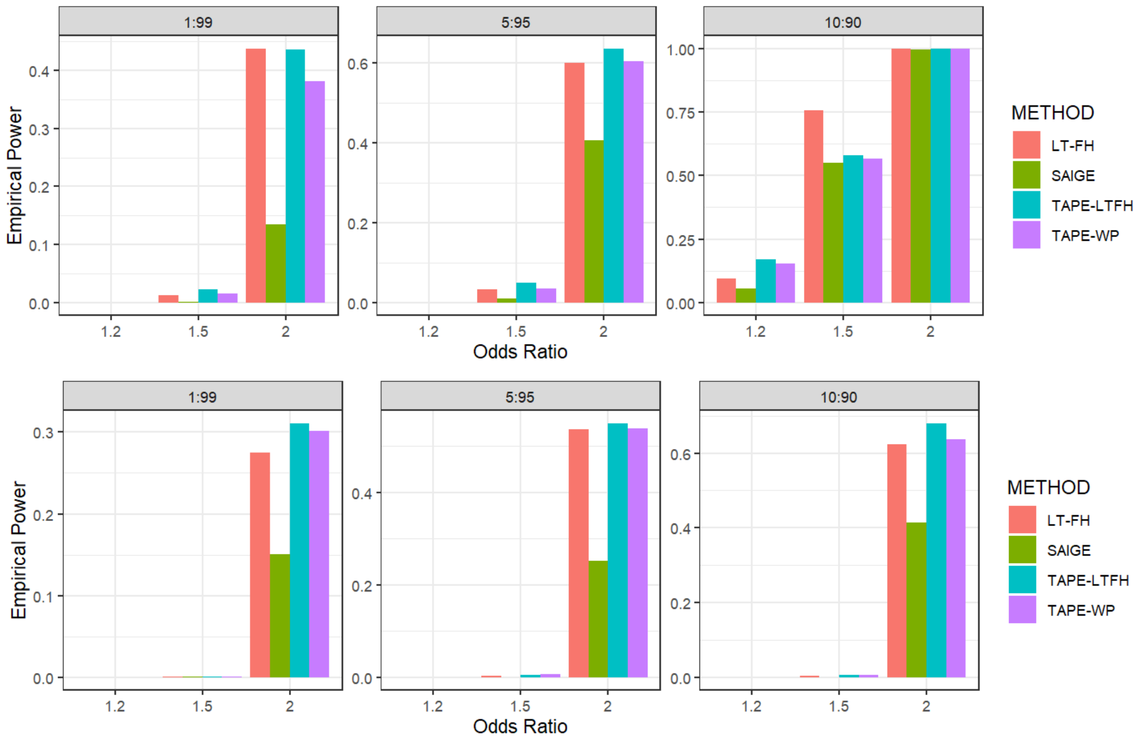

**Supplementary Figure 7.** Log-log plot of computation time under varying sample size, comparing TAPE-WP (labeled as TAPE), SAIGE and LT-FH. Samples were randomly selected from 408,898 white British individuals in UK Biobank data for Type II diabetes using $M = 100{,}000$ variants. Projected computation time for 21 million variants with MAF $\geq 0.01\%$ was plotted. Computation time for TAPE-LTFH is similar to that for TAPE-WP and is therefore omitted.

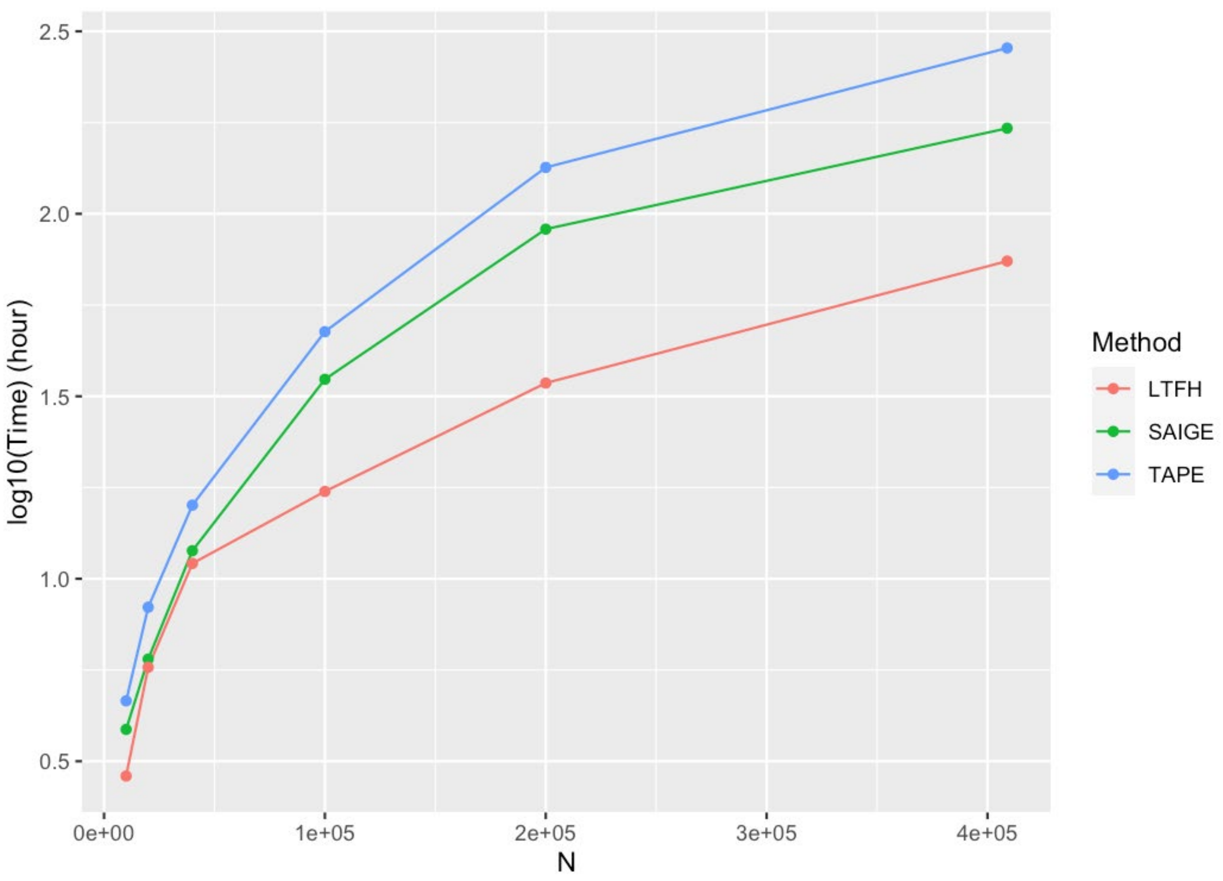

**Supplementary Figure 8.** Q-Q plots for 10 phenotypes in UK Biobank.

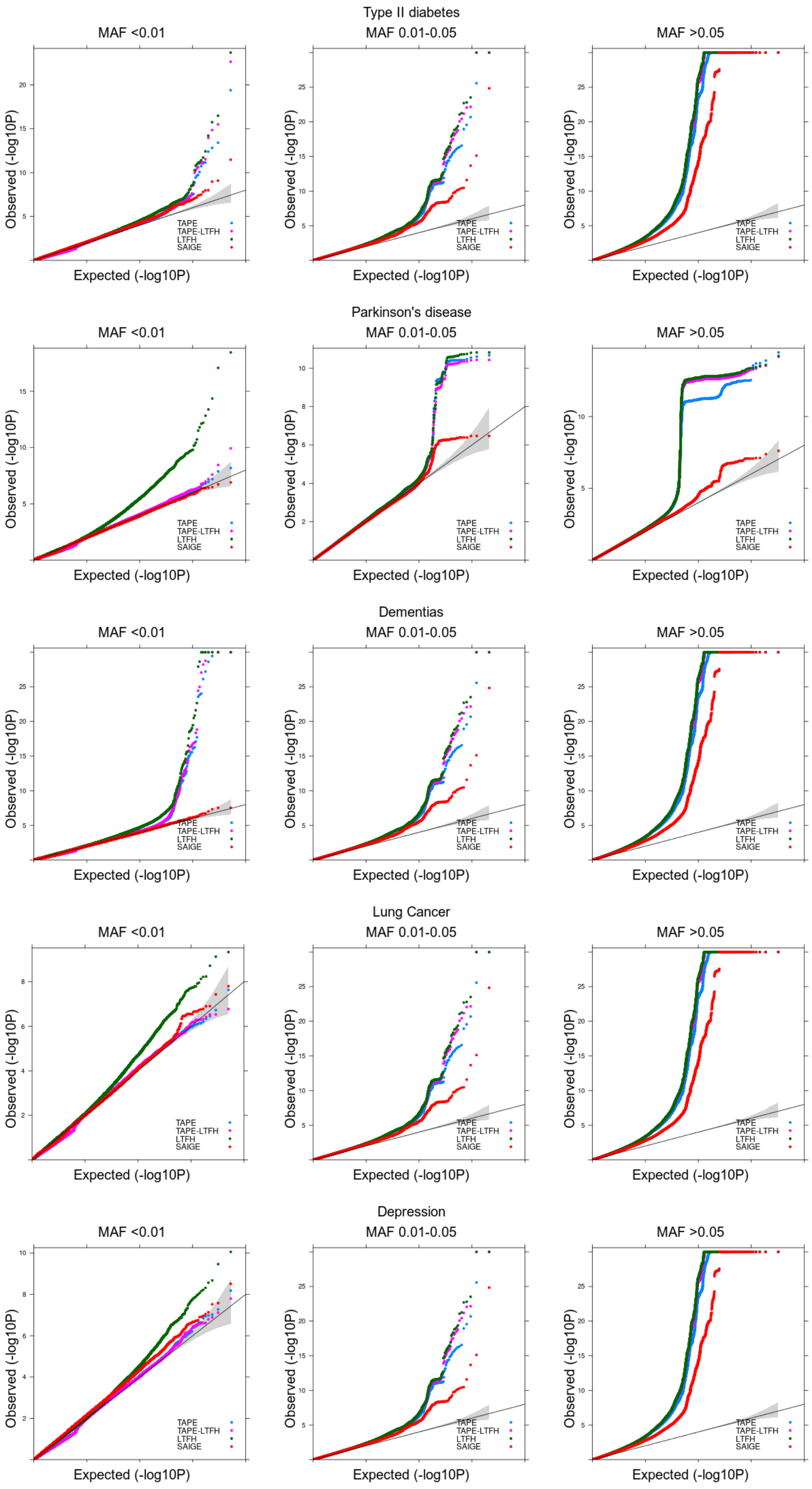

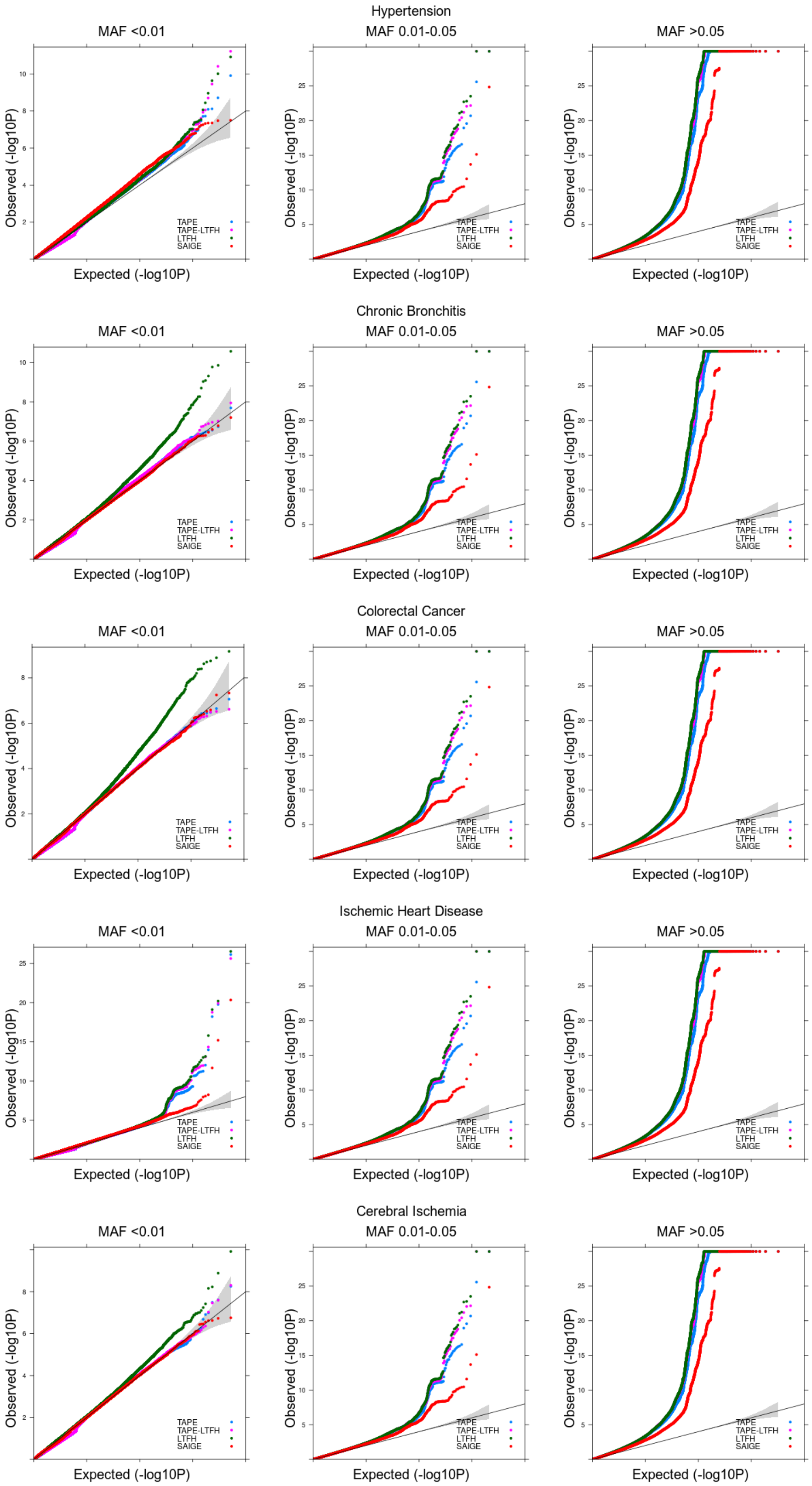
Hypertension
MAF <0.01
MAF 0.01-0.05
MAF >0.05
Chronic Bronchitis
MAF <0.01
MAF 0.01-0.05
MAF >0.05
Colorectal Cancer
MAF <0.01
MAF 0.01-0.05
MAF >0.05
Ischemic Heart Disease
MAF <0.01
MAF 0.01-0.05
MAF >0.05
Cerebral Ischemia
MAF <0.01
MAF 0.01-0.05
MAF >0.05
Observed (-log10P)
Expected (-log10P)
TAPE
TAPE-LTFH
LTFH
SAIGE

**Supplementary Figure 9.** Manhattan plots for 10 phenotypes in UK Biobank. Red: clumped significant variants from TAPE that were not detected by SAIGE; blue: clumped significant variants detected by both TAPE and SAIGE. **a:** results from TAPE-WP; **b:** results from TAPE-LTFH.

a.

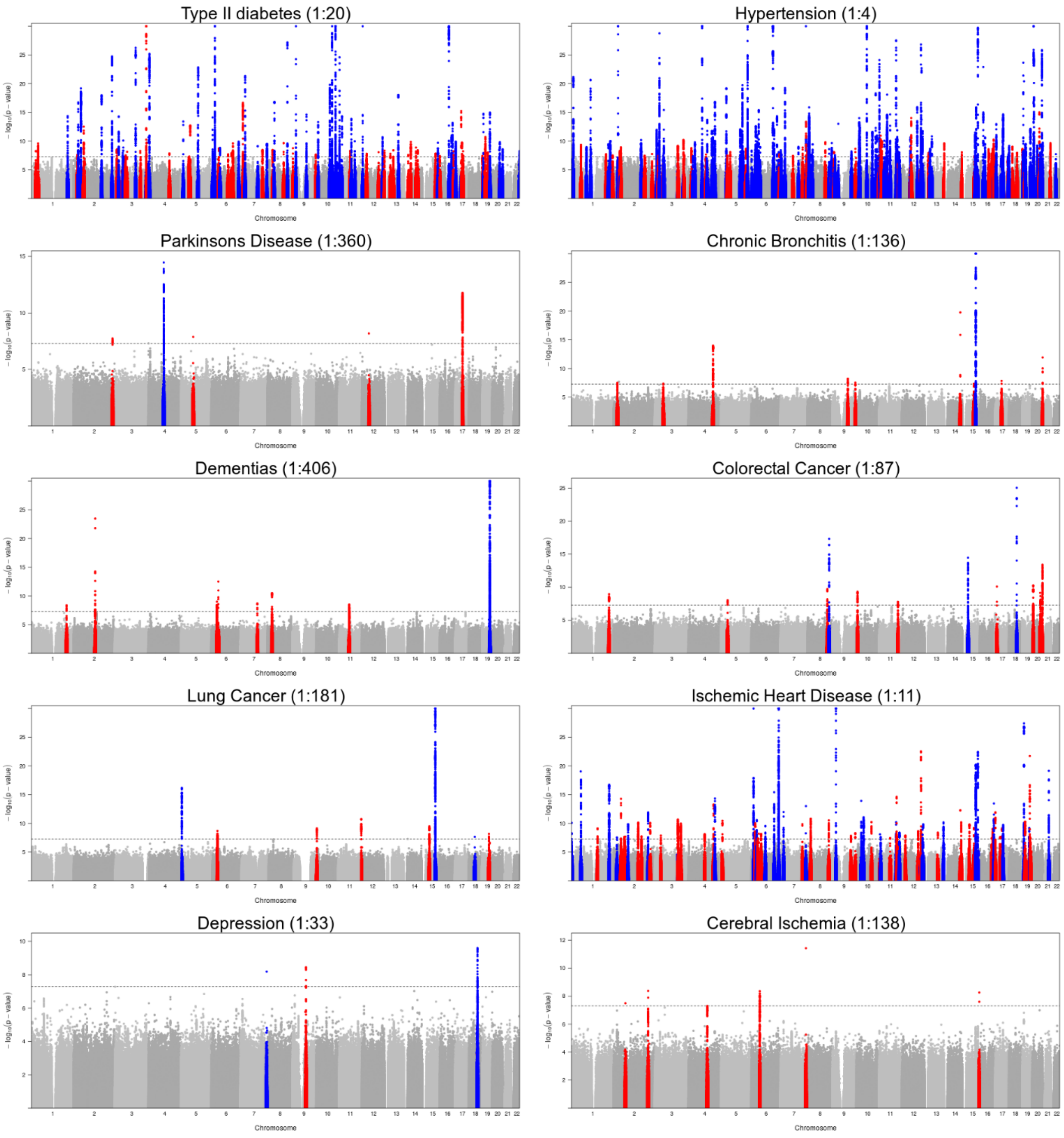

b.

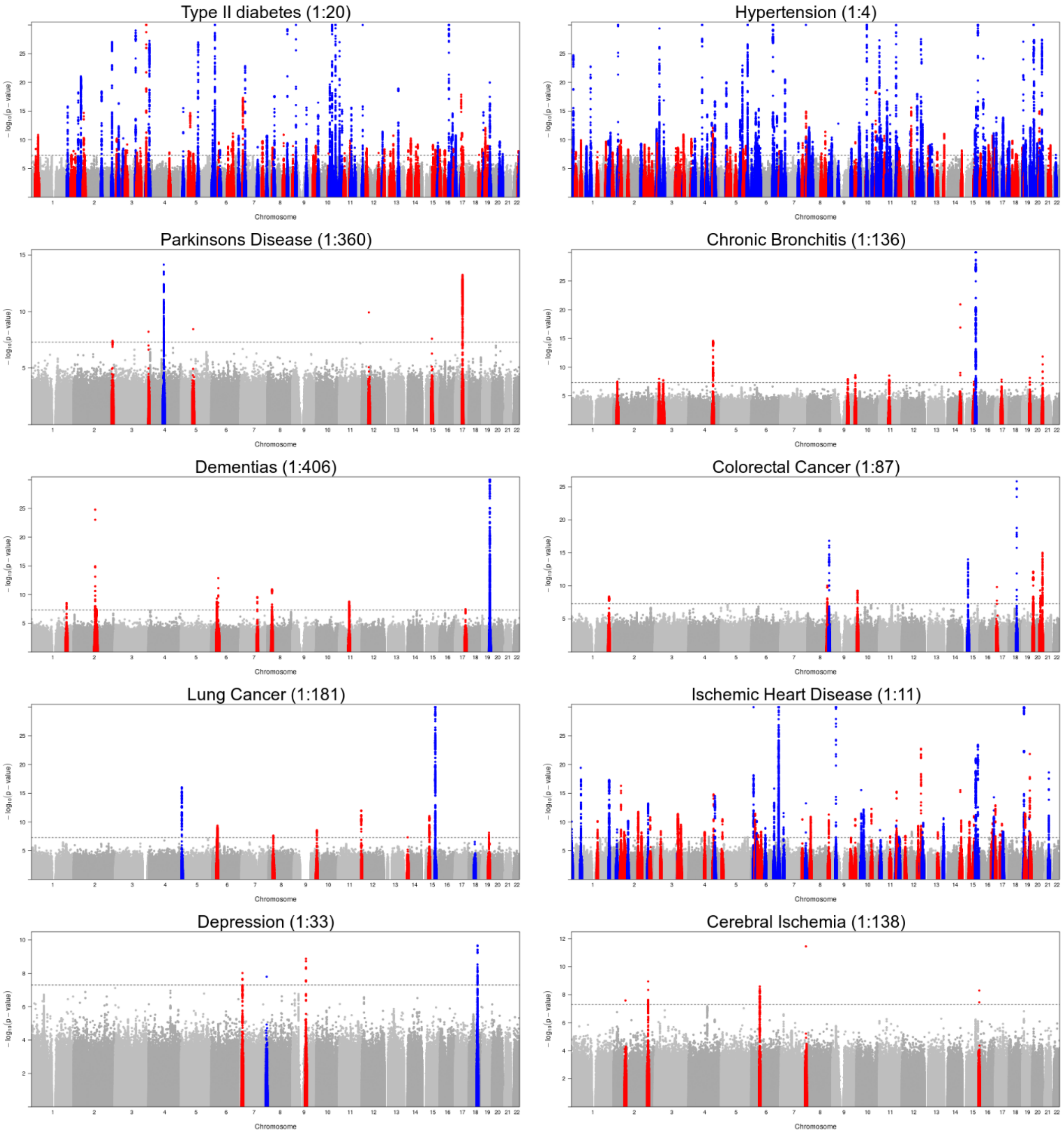
Type II diabetes (1:20)
Hypertension (1:4)
Parkinsons Disease (1:360)
Chronic Bronchitis (1:136)
Dementias (1:406)
Colorectal Cancer (1:87)
Lung Cancer (1:181)
Ischemic Heart Disease (1:11)
Depression (1:33)
Cerebral Ischemia (1:138)
Chromosome

**Supplementary Figure 10.** Manhattan plot for the KoGES data association test results from SAIGE (first row), TAPE-LTFH (second row) and TAPE-WP (third row) (N=72,298). **a**: Diabetes; **b**: Gastric Cancer. For plots from TAPE-WP, red marks clumped significant variants from TAPE-WP that were not detected by SAIGE; blue marks clumped significant variants detected by both TAPE and SAIGE. Significant clumped variants are identified using a window width of 5Mb and a linkage disequilibrium threshold of 0.1.

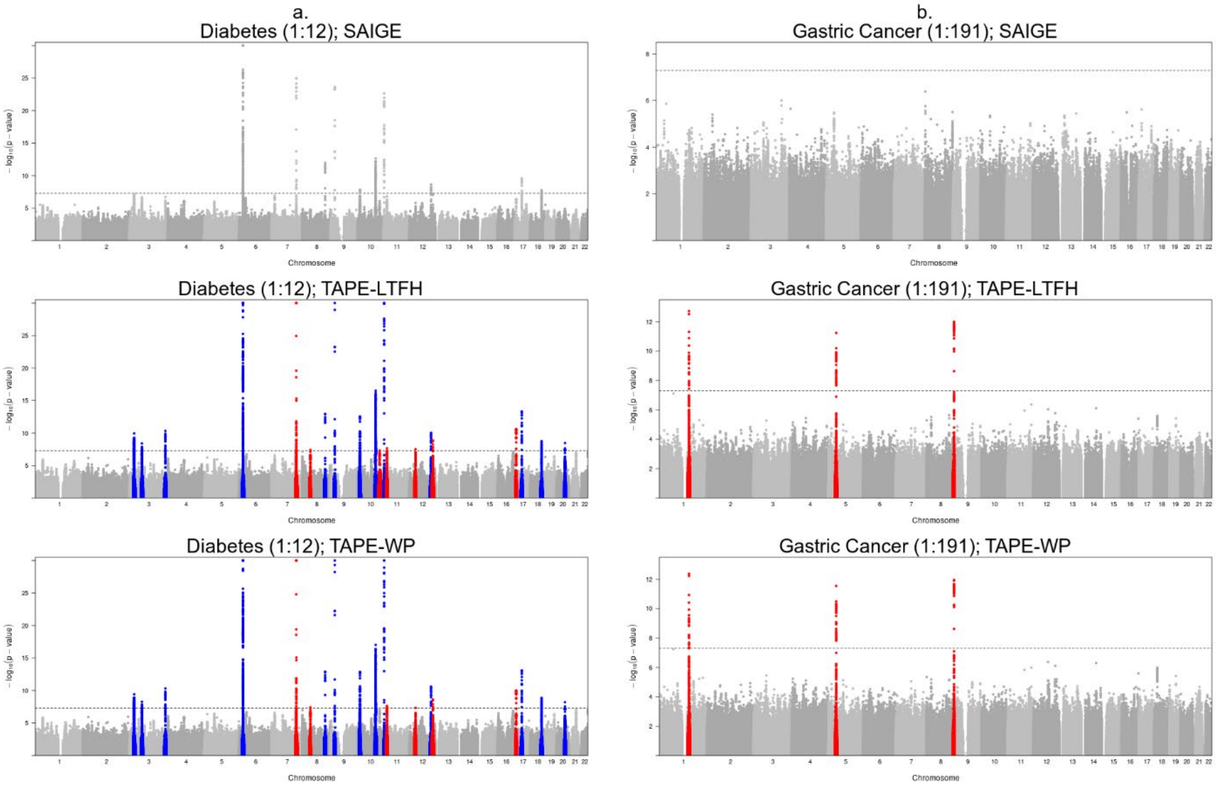

**Supplementary Figure 11.** Q-Q plot for KoGES results from SAIGE, TAPE-LTFH and TAPE-WP (N=72,298), categorized by MAF. Up: Diabetes; Bottom: Gastric Cancer

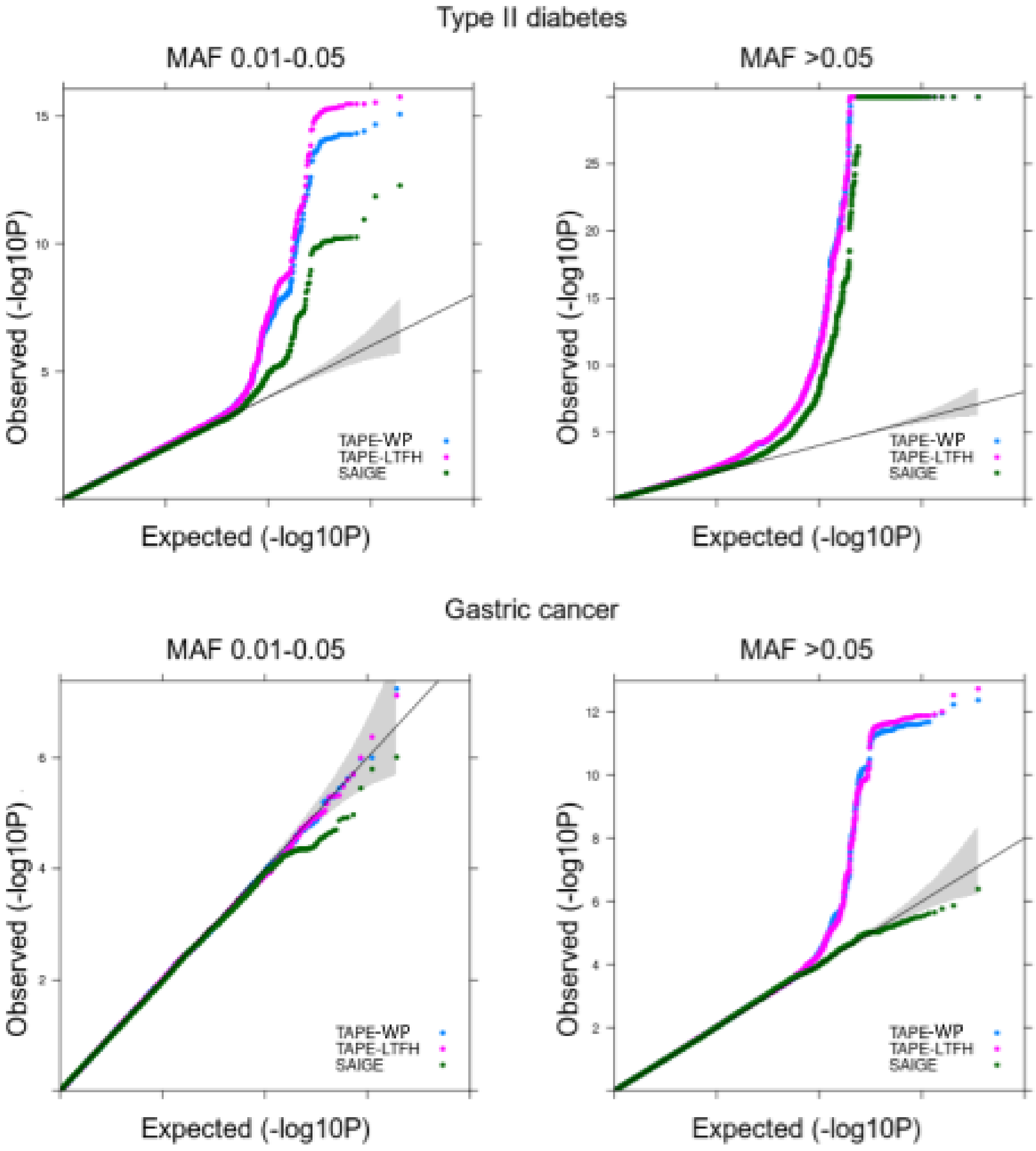

**Supplementary Figure 12.** Scatter plots for 10 phenotypes in UK Biobank. X axis: negative log p-values from SAIGE; Y axis: negative log p-values from TAPE-WP.

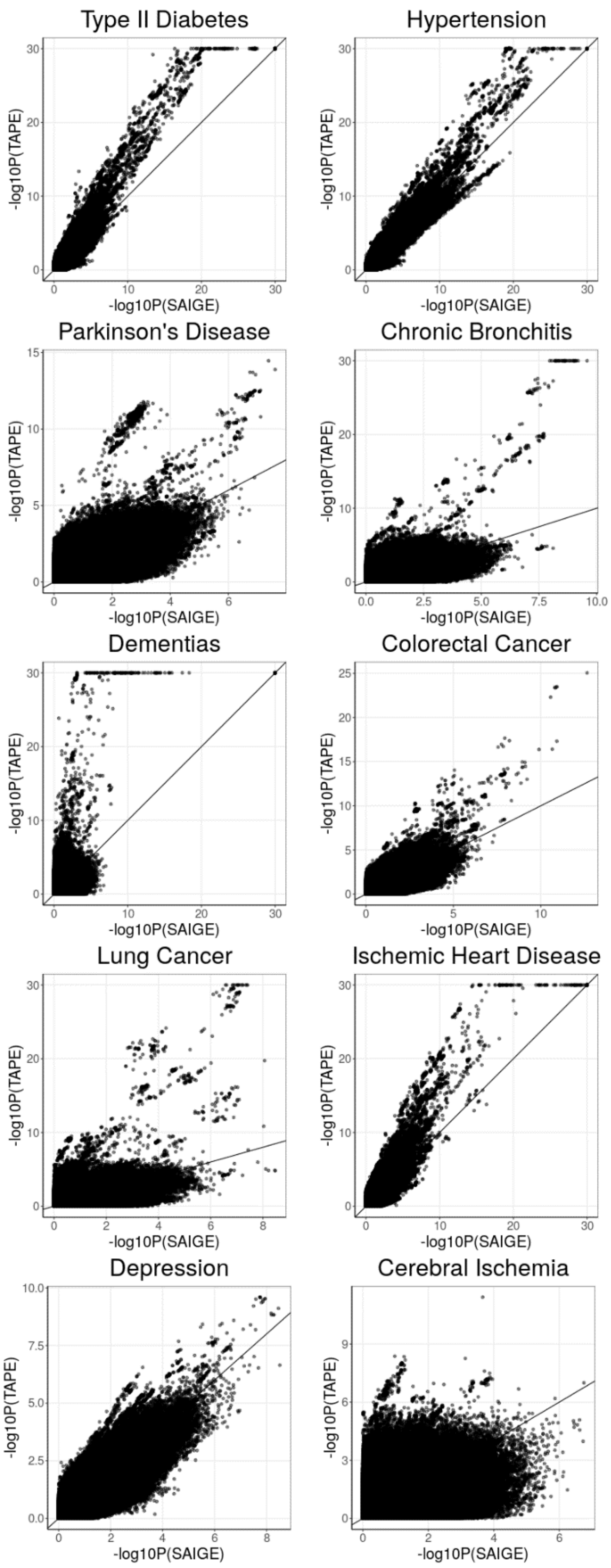

**4. Supplementary Tables**

**Supplementary Table 1.** Number of significant variants and loci for 10 binary traits (case-control ratio in parentheses) in UK Biobank data among white British individuals. **a**: Number of significant variants; **b**: Number of significant clumped variants; **c**: Number of significant clumped variants with MAF < 1%. Significant clumped variants are identified using a window width of 5Mb and a linkage disequilibrium threshold of $r^2 = 0.1$.

**a.**

| Phenotype (case-control ratio) | #Variants (TAPE-WP) | #Variants (TAPE-LTFH) | #Variants (SAIGE) |
|---|---|---|---|
| Type II diabetes (1:20) | 4480 | 5450 | 1737 |
| Parkinson's Disease (1:360) | 2635 | 2628 | 2 |
| Dementias (1:406) | 831 | 892 | 128 |
| Lung Cancer (1:181) | 757 | 885 | 11 |
| Depression (1:33) | 59 | 76 | 22 |
| Hypertension (1:4) | 10721 | 11284 | 7246 |
| Chronic Bronchitis (1:136) | 456 | 552 | 81 |
| Colorectal Cancer (1:87) | 493 | 432 | 65 |
| Ischemic Heart Disease (1:11) | 4909 | 5243 | 1799 |
| Cerebral Ischemia (1:138) | 50 | 67 | 0 |

**b.**

| Phenotype (case-control ratio) | #Variants (TAPE-WP) | #Variants (TAPE-LTFH) | #Variants (SAIGE) |
|---|---|---|---|
| Type II diabetes (1:20) | 154 | 181 | 70 |
| Parkinson's Disease (1:360) | 7 | 9 | 1 |
| Dementias (1:406) | 63 | 67 | 14 |
| Lung Cancer (1:181) | 11 | 14 | 3 |
| Depression (1:33) | 3 | 4 | 3 |
| Hypertension (1:4) | 252 | 273 | 185 |
| Chronic Bronchitis (1:136) | 12 | 15 | 2 |
| Colorectal Cancer (1:87) | 16 | 16 | 3 |
| Ischemic Heart Disease (1:11) | 139 | 142 | 63 |
| Cerebral Ischemia (1:138) | 6 | 5 | 0 |

**c.**

| Phenotype (case-control ratio) | #Variants (TAPE-WP) | #Variants (TAPE-LTFH) | #Variants (SAIGE) |
|---|---|---|---|
| Type II diabetes (1:20) | 8 | 17 | 9 |
| Parkinson's Disease (1:360) | 2 | 2 | 0 |
| Dementias (1:406) | 11 | 12 | 2 |
| Lung Cancer (1:181) | 0 | 3 | 0 |
| Depression (1:33) | 2 | 2 | 2 |
| Hypertension (1:4) | 2 | 10 | 8 |
| Chronic Bronchitis (1:136) | 0 | 4 | 0 |
| Colorectal Cancer (1:87) | 0 | 0 | 0 |
| Ischemic Heart Disease (1:11) | 8 | 13 | 4 |
| Cerebral Ischemia (1:138) | 0 | 0 | 0 |

**Supplementary Table 2.** Attenuation ratios from TAPE-WP, TAPE-LTFH and LT-FH for 10 binary traits (case-control ratio in parentheses) in UK Biobank data among white British individuals.

| | TAPE-WP | TAPE-LTFH | LT-FH |
|---|---|---|---|
| Type II diabetes (1:20) | 0.1092 | 0.1208 | 0.1418 |
| Parkinson's Disease (1:360) | 0.1253 | 0.2220 | 0.4615 |
| Dementias (1:406) | 0.3228 | 0.3491 | 0.5508 |
| Lung Cancer (1:181) | 0.1322 | 0.1814 | 0.2715 |
| Depression (1:33) | 0.0000 | 0.0414 | 0.1147 |
| Hypertension (1:4) | 0.0706 | 0.0752 | 0.0875 |
| Chronic Bronchitis (1:136) | 0.1726 | 0.2109 | 0.2558 |
| Colorectal Cancer (1:87) | 0.0320 | 0.0821 | 0.2105 |
| Ischemic Heart Disease (1:11) | 0.0722 | 0.0781 | 0.1102 |
| Cerebral Ischemia (1:138) | 0.1597 | 0.1752 | 0.3243 |

**Supplementary Table 3.** Computation time (hour) under varying sample size, comparing TAPE-WP (labeled as TAPE), SAIGE and LT-FH. Samples were randomly selected from 408,898 white British individuals in UK Biobank data for Type II diabetes. Null model time: Time (hour) for null model estimation using $M = 100{,}000$ variants was reported. P-value time: Projected computation time (hour) for testing 21 million variants with MAF $\geq 0.01\%$ was listed. Computation time for TAPE-LTFH is similar to that for TAPE-WP and is therefore omitted in the table.

| N | Method | Null Model Time | P-value Time | Total Time |
|---|---|---|---|---|
| 10000 | TAPE | 1.27 | 3.36 | 4.63 |
| 10000 | SAIGE | 2.02 | 1.84 | 3.86 |
| 10000 | LT-FH | 0.15 | 2.73 | 2.88 |
| 20000 | TAPE | 2.84 | 5.51 | 8.35 |
| 20000 | SAIGE | 2.66 | 3.36 | 6.02 |
| 20000 | LT-FH | 0.16 | 5.56 | 5.72 |
| 40000 | TAPE | 5.98 | 9.93 | 15.91 |
| 40000 | SAIGE | 5.28 | 6.65 | 11.93 |
| 40000 | LT-FH | 0.20 | 10.82 | 11.02 |
| 100000 | TAPE | 24.00 | 23.53 | 47.53 |
| 100000 | SAIGE | 19.98 | 15.23 | 35.21 |
| 100000 | LT-FH | 0.85 | 16.49 | 17.34 |
| 200000 | TAPE | 87.78 | 46.21 | 133.99 |
| 200000 | SAIGE | 60.17 | 30.61 | 90.78 |
| 200000 | LT-FH | 2.14 | 32.24 | 34.38 |
| 408898 | TAPE | 190.25 | 94.47 | 284.72 |
| 408898 | SAIGE | 109.02 | 62.58 | 171.60 |
| 408898 | LT-FH | 6.51 | 67.70 | 74.21 |

**Supplementary Table 4.** Disease prevalence of sample individuals and their relatives for two binary phenotypes in the KoGES data.

| Prevalence | Diabetes | Gastric Cancer |
|---|---|---|
| Among samples | 7.6% | 0.52% |
| Paternal | 4.6% | 4.38% |
| Maternal | 8.1% | 2.30% |
| Siblings | 6.3% | 2.51% |
